# Informing the need for a SARS-CoV-2 booster based upon the immune response among young, healthy adults to variants circulating during late 2023

**DOI:** 10.1101/2023.12.16.23300003

**Authors:** Huy C. Nguyen, Kerri G. Lal, Corey A. Balinsky, Lauren Smith, Li Pan, Ying Cheng, Isabella Fox, Robert D. Hontz, Stephen E. Lizewski, Hayley S. Foo, Shelly J. Krebs, Peifang Sun, Andrew G. Letizia

## Abstract

COVID-19 remains a global public health challenge due to ongoing emergence of new immune-evasive SARS-CoV-2 variants, heterogeneous immunity, poor education campaigns, and suboptimal surveillance. In this cross-sectional study, we evaluated the adaptive immune responses in US active-duty service members who completed a COVID-19 primary vaccine series to 3 previously dominant variants (Ancestral, Delta, BA.5) and compared them to 3 variants currently circulating (XBB.1.5, EG.5, and BA.2.86). Analyses were performed based upon time (within or beyond 12 months) and type (vaccine or infection) of most recent exposure. Significant reduction was observed in binding antibodies, neutralization antibodies, memory B cells, and CD8^+^ T cells against current circulating variants compared to previous dominant variants. The reduction in antibody response was more pronounced in those whose most recent exposure, regardless if vaccination or infection, was greater than 12 months from study enrollment. In contrast, the CD4^+^ T cell response was largely consistent across all tested variants. Our study did not show that the type exposure in the last 12 months was a significant factor in determining the magnitude of immune responses. However, the antibody responses to circulating variants were decreased among participants who received the bivalent booster compared to those with an infection in the past 12 months, potentially due to immunological imprinting of the Ancestral strain. Administration of the new XBB.1.5-based booster is likely to enhance cross-reactive humoral immune responses against current SARS-CoV-2 circulating strains.

## BACKGROUND

Coronavirus disease 2019 (COVID-19) remains a leading cause of hospitalizations and deaths world-wide [1] and etiology of long-term sequelae affecting over 18 million Americans [2,3]. The challenge of severe acute respiratory syndrome coronavirus-2 (SARS-CoV-2), the causative agent of COVID-19, is further entrenched due to the continued emergence of variants [4,5] with greater immune escape [6,7]. Durable responses from CD8^+^ T cells, CD4^+^ T cells and memory B cells are known to work in concert with other components of the immune system for long-term virologic control and prevention of severe outcomes [8,9,10]. However, not all components of the cellular immune system contract at the same rate [10] across different demographic groups with varying clinical characteristics [11]. Furthermore, neutralization antibody (NAb) titers to Omicron variants are significantly lower than those to earlier lineages [12,13], and the antibody response wanes over time post-vaccination or post-infection [14,15].

In September 2023, the new monovalent XBB.1.5 booster was recommended for individuals older than 6 months [16] but questions remain regarding its ability to elicit broad immune responses and the need is controversial among less vulnerable populations [17]. Instead, tailored vaccination strategies have been proposed [18]. US active-duty military personnel are a community who face a higher risk of SARS-CoV-2 infections due operating in congregant settings [19], work abroad, and policy changes [20], but in general do not suffer from severe disease. In the face of growing vaccine hesitancy [21]; misinformation [22]; decreased vaccine uptake [23] especially among young adults; and an upcoming winter season with likely rising COVID-19 activity [24], we examine adaptive immunity of US active-duty service members against circulating variants to inform contemporary vaccination recommendations.

## METHODS

### Study Design, Enrollment, and Sampling

A cross-sectional study was conducted over a three-week period in September 2023, comprising 14 enrollment events in Yokosuka, Japan. US Department of Defense active-duty personnel at least 18 years old who completed a COVID-19 vaccine primary series were eligible. Exclusion criteria included those who had cold-like symptoms within the past 30 days; a positive COVID-19 test or vaccine within the past 30 days; pregnant or breastfeeding; taking immunosuppressants or diagnosed with an immunocompromising condition. Ethics review was conducted by the Institutional Review Board, Naval Medical Research Command (NAMRU2.2023.0001) in compliance with all applicable federal regulations governing the protection of human subjects.

Study participants completed a questionnaire (Figure S1) and self-reported vaccination dates. Either a vaccination or natural infection by self-report or identified using electronic medical records constituted an exposure event with prioritization given to the record in case of discordance. Prior positive tests were considered distinct events if at least 90 days apart.

### Peripheral blood mononuclear cells (PBMCs) isolation

PBMCs were isolated using LeucoSep separation using a modified National Institute of Allergy and Infectious Diseases procedure [25].

### Selection of Variants

Six variants were selected for analysis. Three were considered Previous Dominant Variants (PDVs) including the Ancestral, Delta, and Omicron BA.5 strains while three were considered Circulating Variants (CVs) at the time of enrollment, including XBB.1.5, EG.5, and BA.2.86, all of which are currently considered Variants of Interest (VOIs) by the World Health Organization. By mid-November 2023, EG.5, BA.2.86, and XBB.1.5 are the three most prevalent lineages worldwide [26].

### ELISA of Receptor-Binding Domain (RBD) and Spike (S) Protein IgG

All six trimeric S and RBD antigens were diluted in phosphate buffered saline without Ca++ and Mg++, the plates were coated with antigens overnight and quantitative ELISA performed as previously described [13]. Plates were read at OD450nm on plate reader. We subtracted responses against the non-specific binding (PBS-coated wells) during data finalization to yield antigen-specific response. SARS-CoV-2-specific IgG concentrations were calculated using an IgG standard and absorbent densities (OD) were converted to ng/ml by a non-linear regression curve. The limit of detection (LOD) for IgG was 10 ng/ml based on standard pre-pandemic samples.

### Pseudovirus Neutralization Assay

Pseudovirus (PV) was produced using the SARS-CoV-2 Spike gene and the lentivirus derived reporter and packaging plasmids previously described [29]. Spike gene sequences were obtained from the GSAID database, codon optimized for human expression with 19 amino acids removed from the C-terminus and cloned into a pcDNA3.1+ expression vector. PV neutralization assays were performed as previously described [30]. Data were analyzed using Prism GraphPad and nonlinear regression to calculate ND50. The LOD of the assay was 30 corresponding to the starting dilution factor. Refer to Supplemental Methods for more details.

### Memory IgG and Plasmablast B Cells Frequencies

Cryopreserved PBMCs were thawed, washed, and stained for viability. Fc receptors were blocked using normal mouse IgG. After centrifuging and removal of supernatant, cells were stained for the subdomains of the S protein of SARS-CoV-2 using fluorochrome conjugated streptavidin tetramers conjugated to the biotinylated RBDs of the six selected variants, together with a cocktail of phenotyping antibodies. SARS-CoV-2 antigen positive B cells were identified amongst plasmablast populations, defined as CD19+, dump channel/viability dye negative, CD10-, CD20- and CD38^bright,^ CD27+, and memory IgG populations defined as CD19+, dump channel negative, CD10-, CD20+, CD27+ CD21-/+ (Figure S2A, S2B). **“**Responders” within each SARS-CoV-2 antigen tetramer were identified as participants with positive frequencies within either the memory IgG or plasmablast gate above that of a negative control donor run on the same day with study samples to draw gates. Refer to Supplemental Methods for more details.

### T Cells Frequencies

Assay methods for Activation-Induced Markers (AIM) were performed as described previously [31]. Briefly, PBMCs were cultured in the presence of SARS-CoV-2-specific S peptide pools belonging to the six strains. After antigen stimulation, PBMCs were washed and stained with a cocktail of antibodies to identify CD4 and CD8. After staining, cells were directly acquired on a FACS CANTO II with 3-lasers and analyzed with FlowJo software. The gating strategy for AIM cells was drawn relative to the negative and positive controls for each donor as shown in Supplementary Figure S2C. The antigen-specific AIM responses were obtained by subtracting the AIM responses in DMSO cultures. The Stimulation Index (SI) was calculated by dividing the % of AIM+ cells in peptides-stimulated cultures against the % of AIM+ cells in the DMSO control. We calculated the limit of detection as equal to the mean plus 1.7 SD using 6 pre-pandemic samples stimulated with the same antigens above [32]. The limits of detection were 0.05% for both CD4 and CD8. “Responders” were defined as participants with T cell frequencies greater than these cutoffs and the SI was greater than 2, after DMSO background subtraction. Refer to Supplemental Methods for more details.

### Statistics

We used GraphPad Prism version 8.0 for data management and analysis. The Wilcoxon signed-rank test was used for analysis of differences among two groups composing of the same participants. Sub-group analyses were conducted by dividing the cohort based on the timing of the last SARS-CoV-2 exposure event, using 12-month timepoint as cutoff. The Mann-Whitney U test was used for analysis of differences among two independent groups. P values <0.05 were considered significant and applied to all statistical analyses.

## RESULTS

### Cohort Description

A total of 169 participating US servicemembers were enrolled into the study (Table 1). Women comprised 24.3%. The median age was 32, ranging from 18 to 50, with 81.7% younger than 40. A total of 46/169 (27.2%) participants had never received a COVID-19 booster while 50/169 (29.6%) participants had received the bivalent booster containing the Ancestral strain (Wu-1) and the BA.4/BA.5 Omicron variant. None of the participants received the newly approved XBB.1.5-based booster. 67 participants (39.6%) had an exposure event within 12 months of enrollment: 48 (71.6%) a vaccine and 19 (28.4%) with breakthrough infection with confirmed positive test. Conversely, 102 participants (60.4%) had an exposure event more than 12 months from enrollment: 73 (71.6%) a vaccine and 29 (28.4%) with breakthrough infection with confirmed positive test.

**Table 1.**
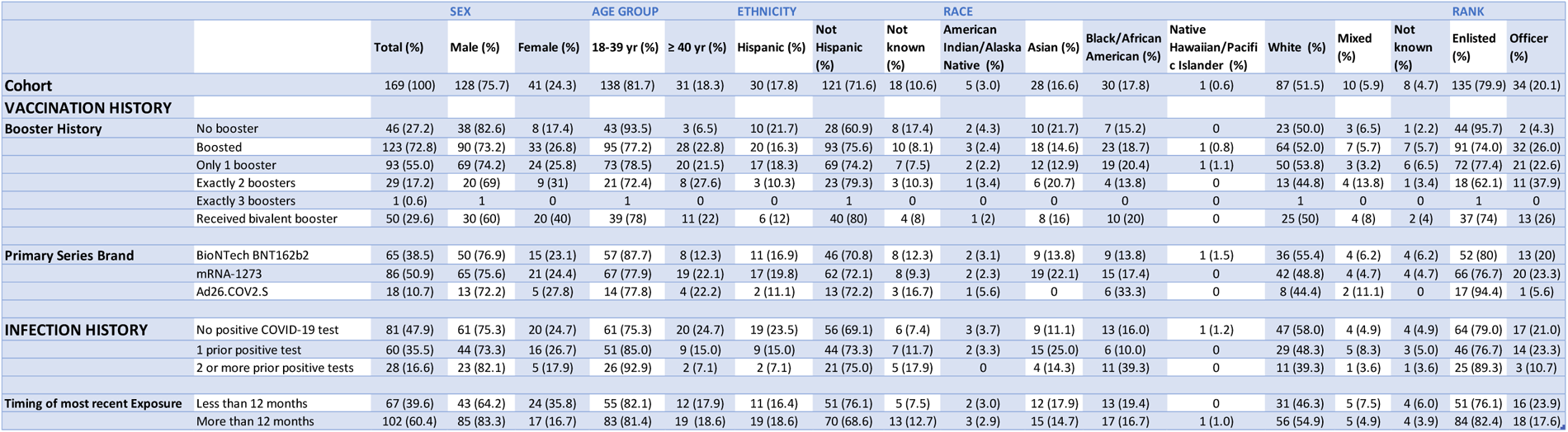
Cohort and exposure groups breakdown by sex, age, ethnicity, race, and rank.

Two participants did not provide a blood sample and seven did not have viable PBMCs post-thaw (Figure S3). Therefore, 167 sera and 160 PBMC samples were analyzed for peripheral immune responses to the previous and current circulating SARS-CoV-2 variants.

### Antibody responses to SARS-CoV-2 were reduced against circulating variants and in participants with exposure greater than 12 months

Immunoglobulin G (IgG) titers to both the RBD and S protein were significantly lower against CVs compared to the Ancestral variant (Figure 1A, 2A). For both RBD and S IgG, the median antibody titers contracted with each new sequential variant with the lowest seen against BA.2.86. Among all tested variants, BA.2.86 had the lowest median titers for both RBD IgG (2,316) and S IgG (14,477) responses.

**Figure 1.**
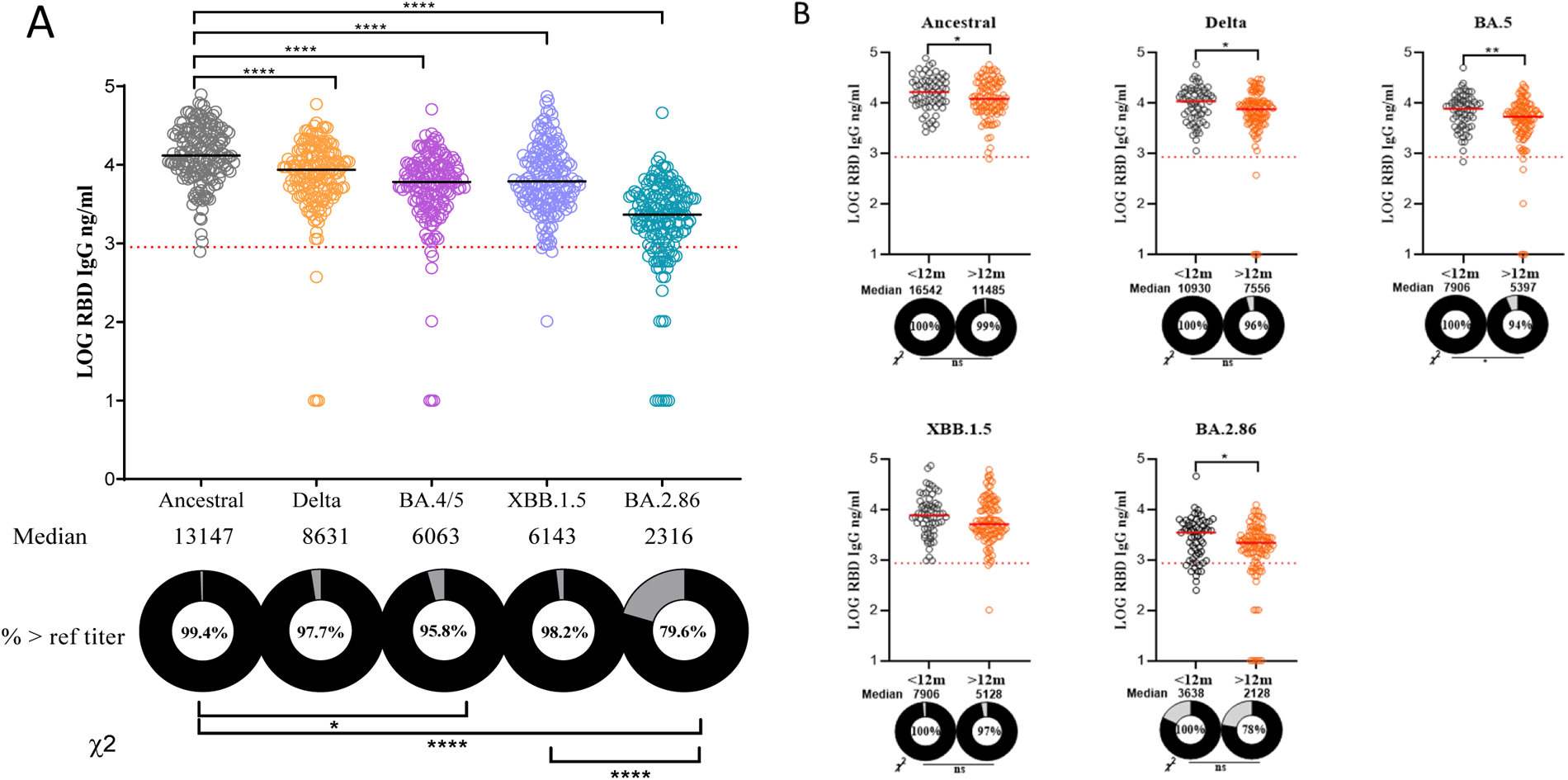
Quantitative IgG antibody to RBD of PDVs and CVs. **A**, variant specific RBD IgG titers for all participants with sera (n = 167). **B**, variant specific RBD IgG titers based on time of most recent exposure (either vaccination or infection) using the 12-month cutoff. Grey circles represent participants with an exposure within 12 months of enrollment while orange circles the participants with an exposure greater than 12 months from enrollment. Log RBD IgG values are shown. EG.5 was not included due to difficulty acquiring EG.5 RBD from vendors. The percentages of participants with titers above the reference “protective” titer (861 ng/ml or 2.94 in log scale) represented as dashed line are shown as donut plots (refer to S. Median titers are shown below each group in ng/ml. Wilcoxon signed-rank test was performed to show group median differences in panel A and Mann-Whitney U test in panel B. Chi square analysis was performed between selected groups. Significant differences are marked as *p < 0.05, **p < 0.01, ***p < 0.001. Abbreviations: IgG, immunoglobulin G; RBD, receptor-binding domain; PDVs, Previous Dominant Variants (Ancestral, Delta, and BA.4/BA.5); CVs, Circulating Variants (XBB.1.5, EG.5, and BA.2.86).

**Figure 2.**
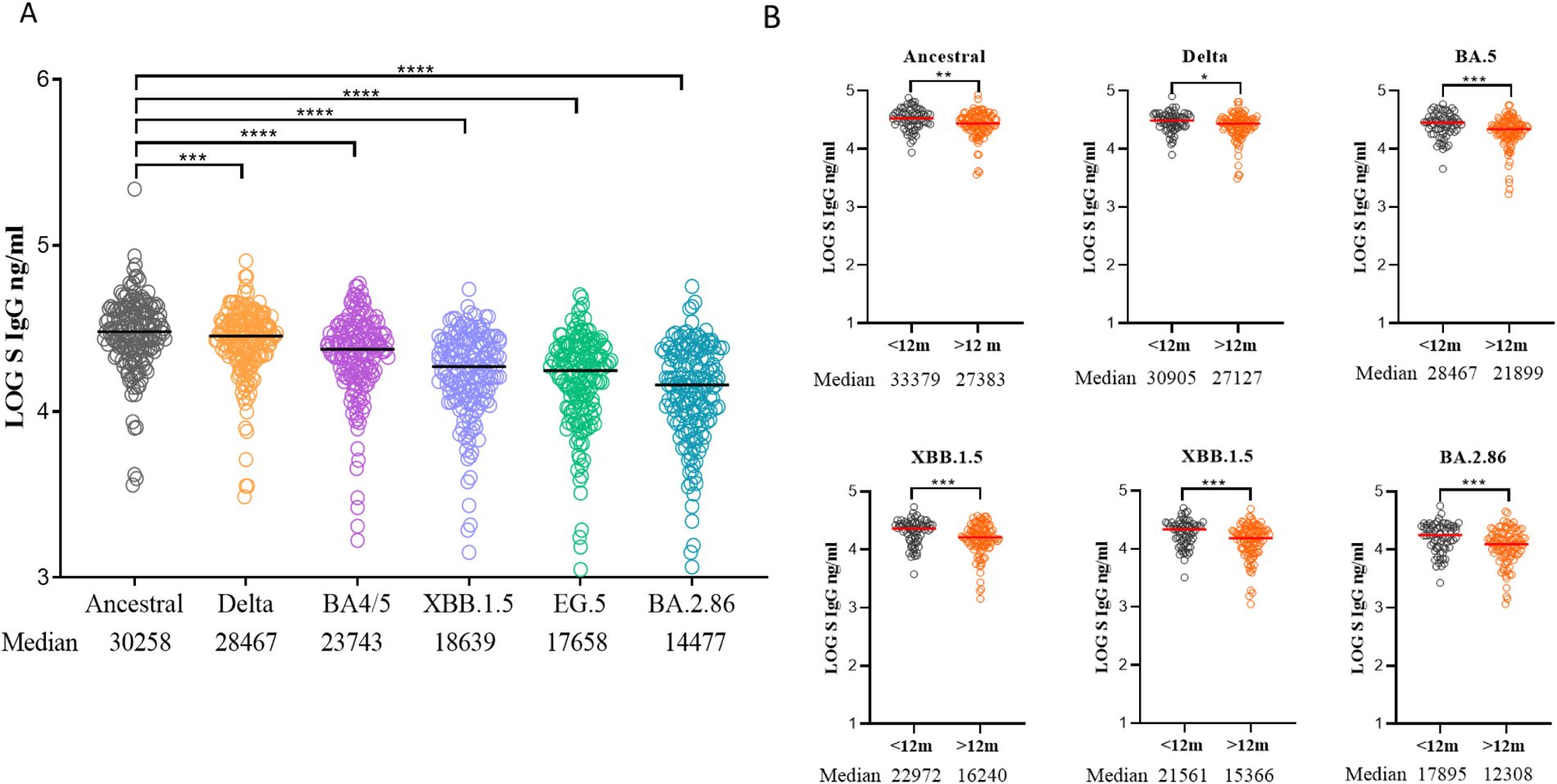
Quantitative IgG antibody to spike (S) protein of PDVs and CVs. **A**, variant specific S IgG titers for all participants with sera (n = 167). **B**, variant specific S IgG titers based on time of most recent exposure (either vaccination or infection) using the 12-month cutoff. Grey circles represent participants with an exposure within 12 months of enrollment while orange circles the participants with an exposure greater than 12 months from enrollment. Log S IgG values are shown. Median titers are shown below each group in ng/ml. Wilcoxon signed-rank test was performed to show group median differences in panel A and Mann-Whitney U test in panel B. Chi square analysis was performed between selected groups. Significant differences are marked as *p < 0.05, **p < 0.01, ***p < 0.001. Abbreviations: IgG, immunoglobulin G; PDVs, Previous Dominant Variants (Ancestral, Delta, and BA.4/BA.5); CVs, Circulating Variants (XBB.1.5, EG.5, and BA.2.86).

Neutralization against the variants followed the same trends as the binding antibodies to RBD and S protein (Figure 3A). NAb titers against all PDVs were higher compared to CVs. EG.5 had the lowest median titer and proportion of donors above LOD (82.6%) among the CVs, followed by XBB.1.5 (83.8%) and BA.2.86 (89.2%), respectively. For PDVs, 1-4% of participants had undetectable NAb activity; for CVs, 11-17% had no detectable NAb response.

**Figure 3.**
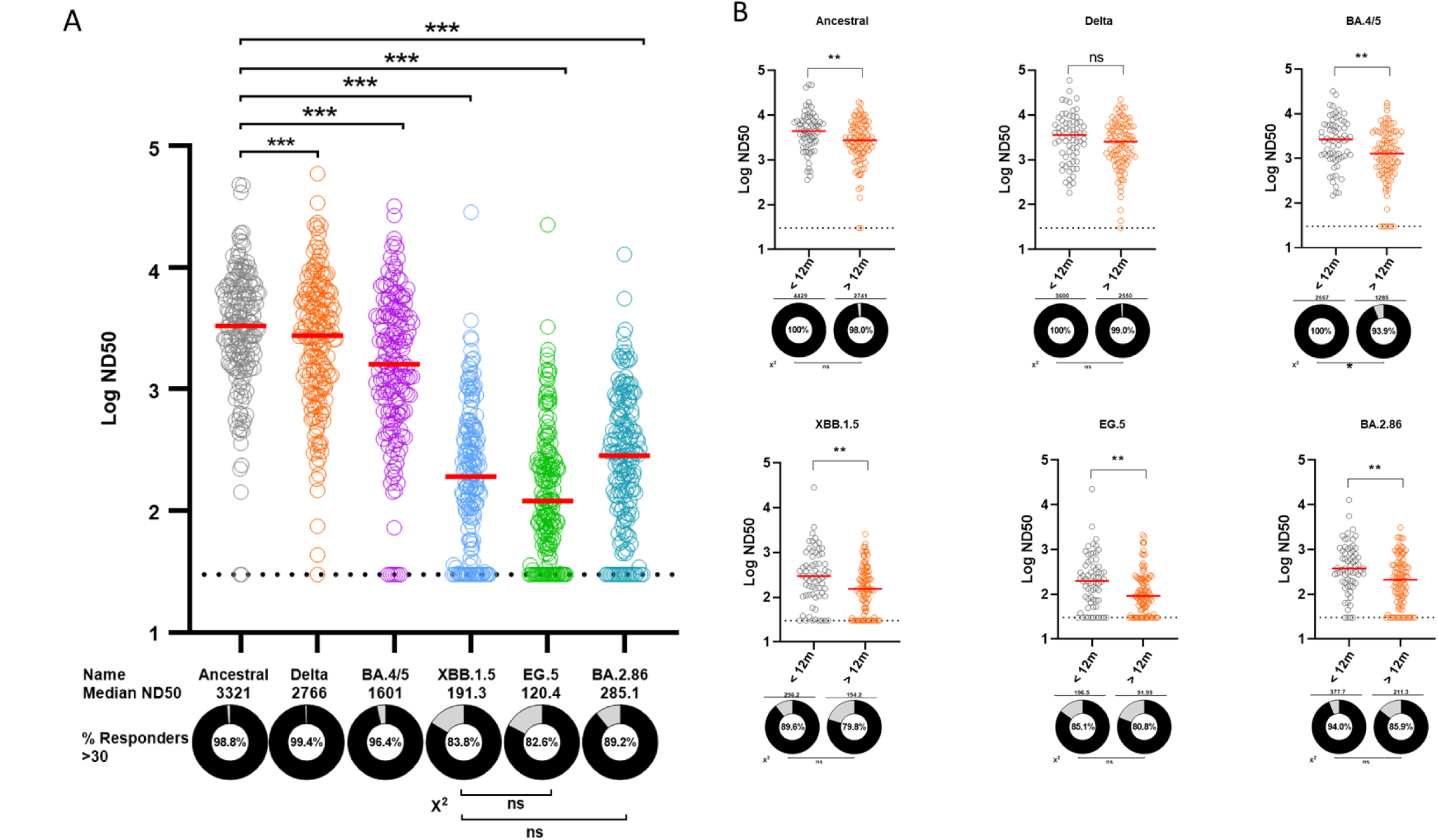
Pseudovirus neutralization antibody (NAb) to RBD of PDVs and CVs. **A**, variant specific NAb titers for all participants with sera (n = 167). **B**, variant specific NAb titers based on time of most recent exposure (either vaccination or infection) using the 12-month cutoff. Grey circles represent participants with an exposure within 12 months of enrollment while orange circles the participants with an exposure greater than 12 months from enrollment. Log ND50 values are shown. The percentages of participants with titers above the LOD (30 or 1.5 in log scale) represented as dashed line are shown as donut plots. Median titers are shown below each group in ng/ml. Wilcoxon signed-rank test was performed to show group median differences in panel A and Mann-Whitney U test in panel B. Chi square analysis was performed between selected groups. Significant differences are marked as *p < 0.05, **p < 0.01, ***p < 0.001. Abbreviations: PDVs, Previous Dominant Variants (Ancestral, Delta, and BA.4/BA.5); CVs, Circulating Variants (XBB.1.5, EG.5, and BA.2.86); LOD, limit of detection.

In addition, for RBD IgG and NAb measured against all tested variants, significantly fewer donors with titters above the LOD or reference “protective” titer were observed in participants whose most recent exposure was greater than 12 months from enrollment (Figure 1B, 3B). Notably, for BA.2.86, while 94% of donors with an exposure in the past 12 months had detectable NAb activity, the proportion of donors above the LOD was only 85.9% among those with an exposure greater than 12 months from enrollment. Overall, for all variants, there was no statistically significant difference in the RBD IgG, S IgG, and NAb titers of participants whose most recent exposure was a vaccine compared to those whose most recent exposure was an infection, regardless if the exposure occurred less than or greater than 12 months from enrollment (exception: RBD IgG to XBB.1.5 in those with an exposure in the last 12 months) (Figure S4, S5).

### B cell responses to SARS-CoV-2 were reduced against circulating variants but largely unaffected by timing and nature of most recent exposure

The frequency of variant RBD specific IgG-positive memory B cells (MBC) against CVs were lower than those against PDVs, with statistical significance reached when comparing the Ancestral strain to all Omicron strains (Figure 4A). The proportion of “responders” was lowest for BA.2.86, followed by XBB.1.5 and EG.5, respectively.

**Figure 4.**
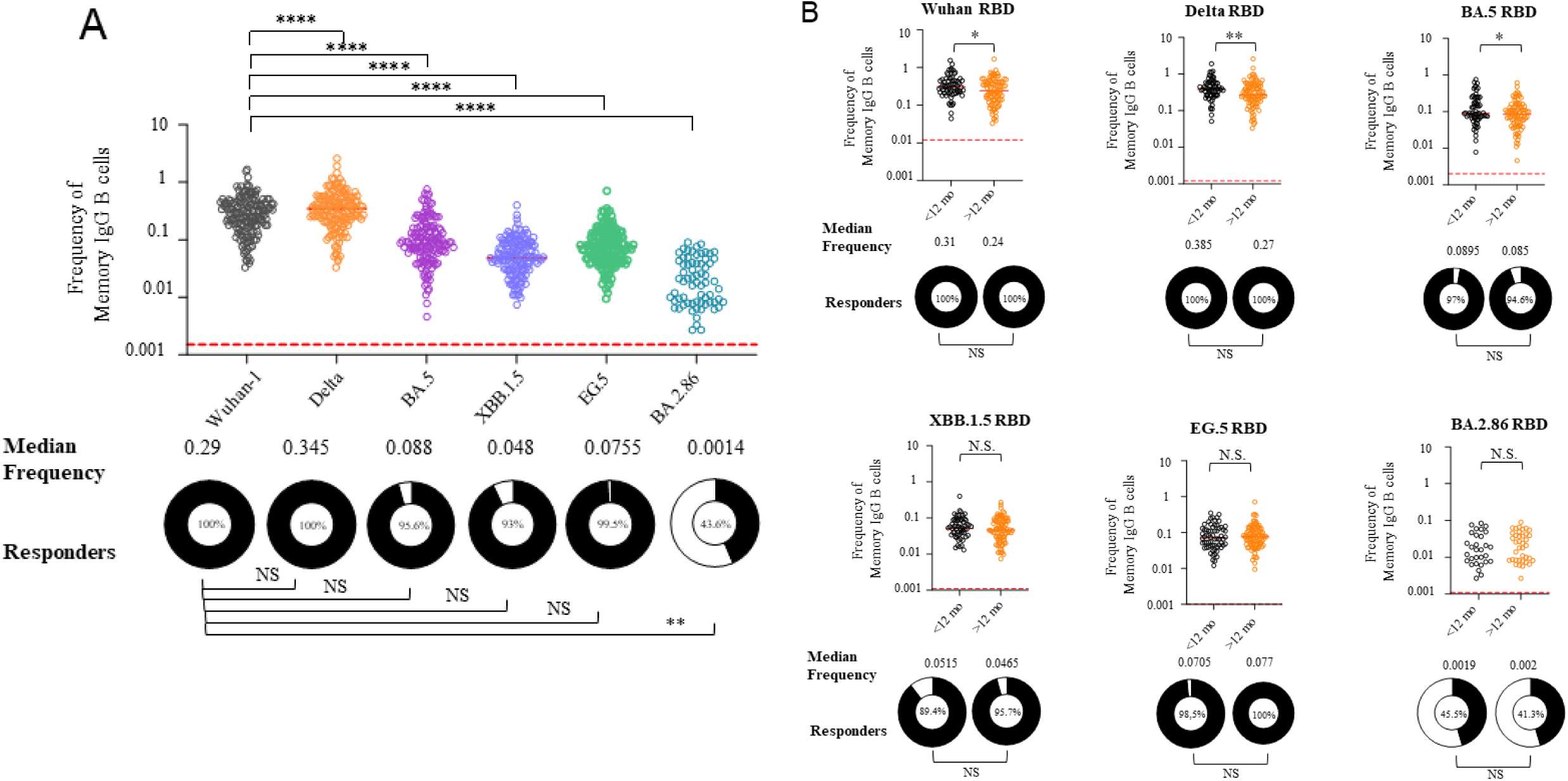
Frequency of IgG positive memory B cells (MBCs) to RBD of PDVs and CVs. **A**, variant specific MBC frequencies for all participants with PBMCs (n = 160). **B**, variant specific MBC frequencies based on time (12-month cutoff) and nature of most recent exposure (either vaccine or infection). Grey circles and squares represent participants with an exposure within 12 months of enrollment while orange circles and squares the participants with an exposure greater than 12 months from enrollment. The percentages of MBC “responders” are shown as donut plots. MBC “responder” is defined as having an MBC frequency higher than the negative control sample run on the same day. The dashed line represents the median of the frequencies of MBCs meeting gating criteria per variant across all days of analysis from a negative control donor. Median frequencies are shown below each group. Wilcoxon signed-rank test was performed to show group median differences in panel A and Mann-Whitney U test in panel B. Significant differences are marked as *p < 0.05, **p < 0.01, ***p < 0.001, ****p<0.0001. Abbreviations: IgG, immunoglobulin G; PDVs, Previous Dominant Variants (Ancestral, Delta, and BA.4/BA.5); CVs, Circulating Variants (XBB.1.5, EG.5, and BA.2.86); PBMCs, peripheral blood mononuclear cells.

In addition, for all CVs, the MBC frequencies in participants were statistically similar among all participants when using the 12-month cutoff for most recent exposure (Figure 4B). In contrast, for the PDVs, MBC frequencies were statistically higher among those with a more recent exposure (within 12 months of enrollment) compared to those with a more remote exposure (greater than 12 months from enrollment). There was no significant difference in MBC frequencies based upon most recent exposure type regardless of timing (Figure S6).

Assessment of plasmablasts to tested SARS-CoV-2 variants followed a similar trend with higher median frequencies more commonly observed against PDVs compared to CVs (Figure S8). Of note, however, the lowest RBD-bound proportion of memory B cells was observed for the Delta variant, likely due to higher activation and secretion of B cell receptors (BCRs) from B cell surfaces.

### T cell responses to SARS-CoV-2 were varied against circulating variants but largely unaffected by timing and nature of most recent exposure

The frequency of variant RBD specific CD8^+^ T cells against CVs were lower than those against PDVs, with statistical significance reached when comparing the Ancestral strain to all Omicron strains (Figure 5A). The proportion of “responders” was lowest for BA.2.86 (59.1%), followed by EG.5 (62.3%), and XBB.1.5 (64.2%), respectively. In contrast, CD4^+^ T cell frequencies were consistent with no significant difference across all variants with the notable exception of BA.2.86 and EG.5 when compared to the Ancestral strain (Figure 6A). There was no significant difference in the proportion of “responders” when comparing CVs to the Ancestral variant.

**Figure 5.**
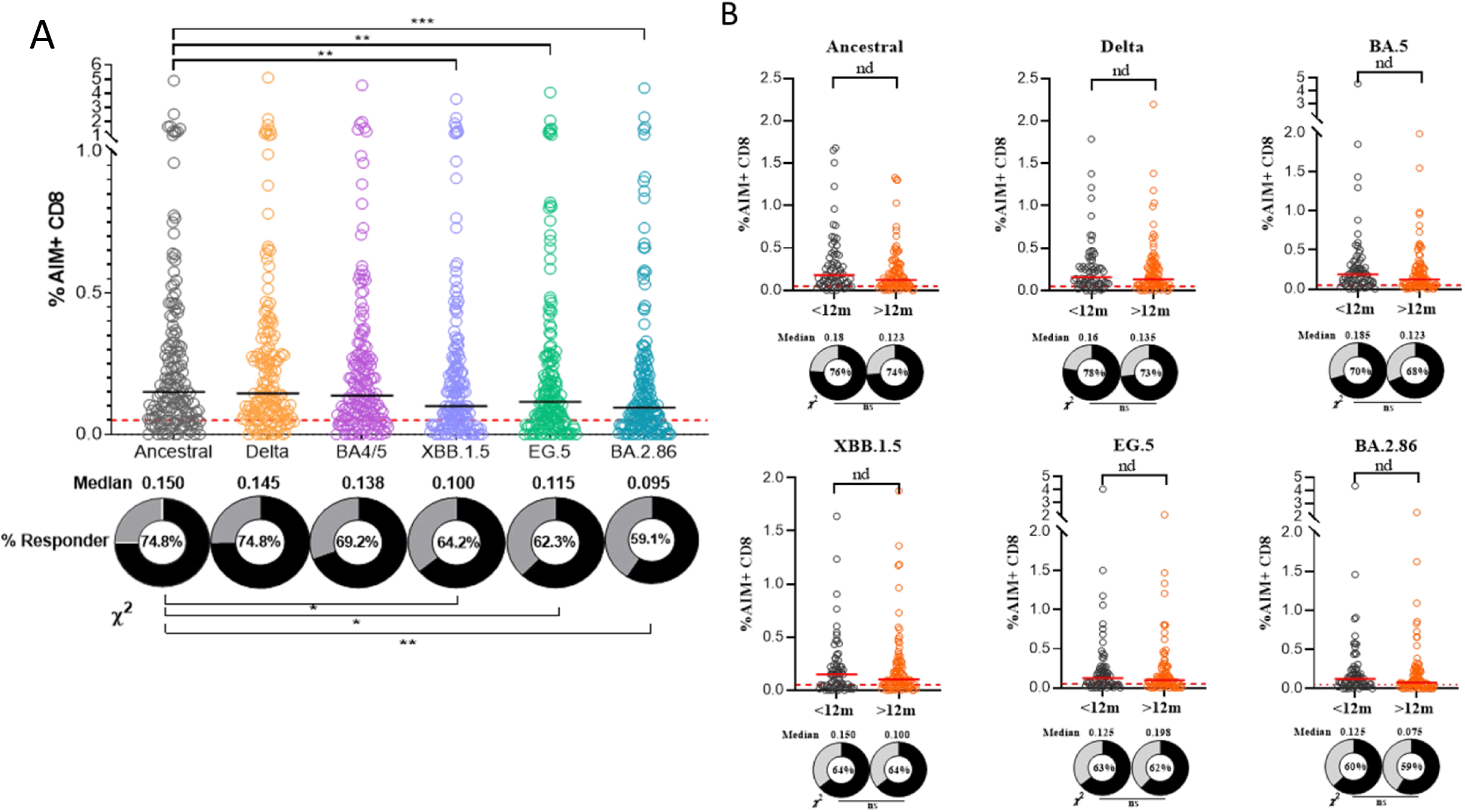
*Frequency of AIM positive CD8+ (cytotoxic) T cell to RBD of* PDVs and CVs. A, variant specific CD8+ T cell frequencies for all participants with PBMCs (n = 160). B, variant specific CD8+ T cell frequencies based on time (12-month cutoff) and nature of most recent exposure (either vaccine or infection). Grey circles and squares represent participants with an exposure within 12 months of enrollment while orange circles and squares the participants with an exposure greater than 12 months from enrollment. AIM positivity defined as CD69+ and CD137+. The percentages of CD8+ T cell “responders” are shown as donut plots. CD8+ T cell “responder” is defined as having a frequency greater than 0.05% and stimulation index greater than 2 (refer to Methods). Median frequencies are shown below each group. Wilcoxon signed-rank test was performed to show group median differences in panel A and Mann-Whitney U test in panel B. Significant differences are marked as *p < 0.05, **p < 0.01, ***p < 0.001. Abbreviations: AIM, activation-induced markers; PDVs, Previous Dominant Variants (Ancestral, Delta, and BA.4/BA.5); CVs, Circulating Variants (XBB.1.5, EG.5, and BA.2.86); PBMCs, peripheral blood mononuclear cells.

**Figure 6.**
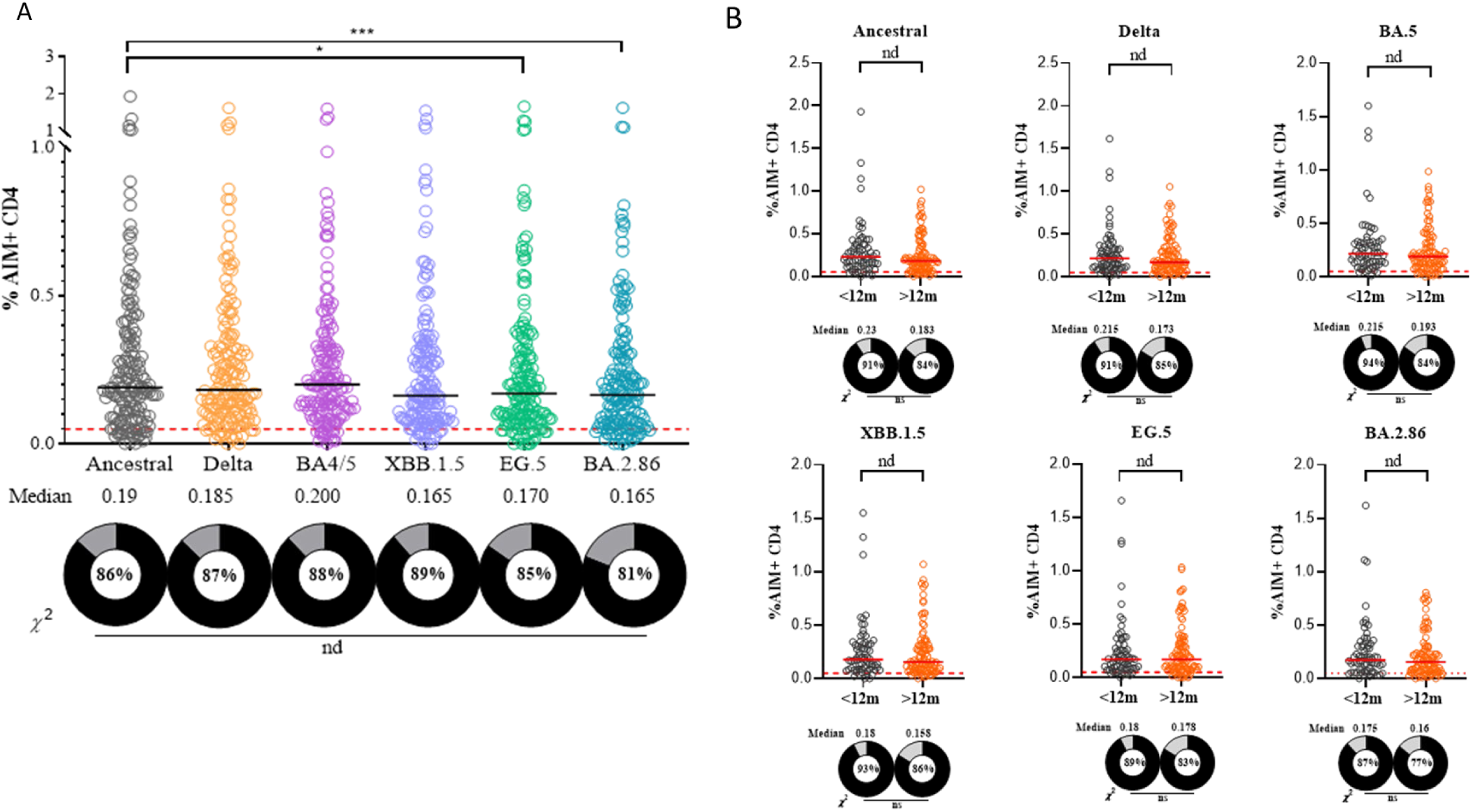
Frequency of AIM positive CD4+ (helper) T cell to RBD of PDVs and CVs. A, variant specific CD4+ T cell frequencies for all participants with PBMCs (n = 160). B, variant specific CD4+ T cell frequencies based on time (12-month cutoff) and nature of most recent exposure (either vaccine or infection). Grey circles and squares represent participants with an exposure within 12 months of enrollment while orange circles and squares the participants with an exposure greater than 12 months from enrollment. AIM positivity defined as CD134+ and CD137+. The percentages of CD4+ T cell “responders” are shown as donut plots. CD4+ T cell “responder” is defined as having a frequency greater than 0.05% and stimulation index greater than 2 (refer to Methods). Median frequencies are shown below each group. Wilcoxon signed-rank test was performed to show group median differences in panel A and Mann-Whitney U test in panel B. Significant differences are marked as *p < 0.05, **p < 0.01, ***p < 0.001. Abbreviations: AIM, activation-induced markers; PDVs, Previous Dominant Variants (Ancestral, Delta, and BA.4/BA.5); CVs, Circulating Variants (XBB.1.5, EG.5, and BA.2.86); PBMCs, peripheral blood mononuclear cells.

For all tested variants, there was no statistically significant difference in CD8^+^ or CD4^+^ T cell response among participants regardless of the timing (Figure 5B, 6B) or nature (Figure S7, S9) of the most recent exposure.

## DISCUSSION

In our young healthy cohort of servicemembers sampled in September 2023, we observed significant reduction in adaptive immune responses including antibodies, neutralization antibodies, memory B cells, and CD8^+^ T cells against CVs compared to PDVs, with the notable exception of CD4^+^ T cells, whose response was detectable and largely consistent for all tested variants. In addition, we noted that a more recent exposure in the last 12 months from either vaccination or infection increased binding and functional antibody responses compared to individuals whose most recent exposure was over 12 months prior to enrollment. Taken together, these results suggest that a booster may enhance humoral and select cellular responses, especially in those without SARS-CoV-2 exposure event in the last 12 months.

We showed that the variant specific RBD and S antibody titers progressively contracted with each chronologically sequential variant starting with the Ancestral strain. Among the CVs, BA.2.86 exhibited the largest fold-reduction in median antibody titers compared to the Ancestral strain, likely reflecting the high number of mutations in BA.2.86 [33]. Similarly, while only 1-4% of participants had undetectable NAb titers against PDVs; the percentage below the limit of detection increased to 11-17% against CVs.

Timing of most recent exposure appeared to have a significant impact on both binding and neutralizing antibody titers. RBD IgG, S IgG, and ND50 titers of those participants with an infection in the past 12 months were significantly higher than those participants with an exposure more than 12 months ago, suggesting an exposure within a year would improve detectable neutralization. Given the high prevalence of XBB.1.5 and its descendant lineages toward the end of 2022 and most of 2023, participants who had a natural infection within 12 months of enrollment in September 2023 were likely infected with the XBB.1.5-lineage strains. An XBB.1.5-based booster, especially for those who lack an exposure in the last year, would likely increase RBD IgG, S IgG, and ND50 titers and enhance the response especially among the participants who lacked neutralizing antibodies to CVs. These results help identify and quantify a minimum number of young and healthy adults that could expect to benefit from the new XBB.1.5-based booster through enhanced antibody responses against CVs.

In parallel with antibody responses, the frequency of peripheral variant RBD-specific memory B cells contracted with each variant, with the lowest frequency observed for BA.2.86. The presence of memory B cells to variant RBDs indicated that these B cells are ready to be recalled upon subsequent exposure to CVs. However, only 43.6% of participants contained BA.2.86-specific memory B cells compared to 90% or higher proportions observed against other CVs. Given that SARS-CoV-2 boosters have been shown to induce de novo B cell responses targeting variant-specific epitopes [34], our results suggest that there is a substantial proportion of individuals who could benefit from the new XBB.1.5-based vaccine beyond antibody responses.

In contrast to antibody responses, no significant difference in CD4^+^ and CD8^+^ T cell responses were observed between participants whose last exposure was less than or greater than 12 months for all variants. This is likely due to a high proportion of T cell epitopes being conserved and therefore unaffected by Omicron mutations combined with the long-term durability of T cell responses following an exposure event [35]. These findings suggest a more expansive and durable response from the cellular immune components compared to humoral counterparts.

Clinically, we would expect the presence of greater quantities of binding and neutralizing antibodies would likely decrease infectivity [36], subsequently lowering the number of missed workdays and risk of transmission to vulnerable populations. The largely consistent CD4^+^ and CD8^+^ T cell responses among circulating variants regardless of the nature of exposure or timing within the last 12 months imply that the new XBB.1.5-based booster may not significantly enhance the consistent CD4^+^ T cell responses already present nor significantly expand the memory T cell pools [37].

In general, the type of most recent exposure, whether vaccine or natural infection, did not have a statistically significant impact on the adaptive immune responses regardless of variant. However, participants who had a natural infection within 12 months of enrollment consistently had significantly higher binding and neutralizing antibody titers against CVs than those who received a vaccine in the past 12 months. Out of the 48 participants who received a vaccine within 12 months, 46 (95.8%) received the bivalent booster that consisted of both the Ancestral and BA.4/BA.5 spike proteins. Our data also showed that exposure within the past 12 months recalled memory B cell responses and, to a lesser degree, T cell responses that were either specific or cross-reactive to PDVs, potentially imprinted upon through prior receipt of early vaccines or previous infections. These findings suggest that the Ancestral component of the bivalent booster, likely via deep immunological imprinting, disadvantageously enhanced immune responses against PDVs while hindering a broader immune response to CVs, consistent with recent reports [38,39,40]. Future COVID-19 vaccines should not contain previously dominant variants no longer in circulation.

This study has several limitations. It is a cross-sectional design collecting data only at one timepoint, limiting an assessment of immune response kinetics. The sample size is relatively small and mostly men, limiting certain sub-group analyses. Only reported cases were considered an exposure, therefore asymptomatic as well as unrecalled and undocumented pauci-symptomatic cases were not identified. None of the participants were evaluated for an active infection: measuring immune responses was compartmentalized and performed ex-vivo under artificial conditions.

The results from this study, even though obtained only from US Navy Sailors, are generalizable to other young and healthy adults. Future studies should examine the immune responses of participants who have received the XBB.1.5-based booster as well as those who only received the primary series. Ongoing SARS-Cov-2 surveillance for immunologic immune escape is also needed.

## CONCLUSIONS

Rapid execution of a comprehensive study investigating variant-specific adaptive immune responses can help inform the need for contemporary SARS-CoV-2 boosters and quantify the potential benefits. The XBB.1.5-based booster would enhance both binding and neutralizing antibody responses to circulating variants, especially those without an exposure in the last 12 months. Continuous surveillance against new variants is needed to inform the need for, composition, and timing of SARS-CoV-2 vaccine boosters.

## Supporting information

References

## Data Availability

All data produced in the present study are available upon reasonable request to the authors.

## Acknowledgements

The authors acknowledge the outstanding support from the Vysnova study team members (Hoa Nguyen, Emily Nguyen, Nikki Mahmood, Mary Ann Serote, Jamie Martin, Gloria Reyes, Genovieve Ambulo, Rhea Ramos, Kimberly Ibanez, and Long Tran) in Japan as well as HM1 Jin Lin, LS2 Matthew Beye, and LCDR Sarah Jenkins who supported the investigators in all aspects of study especially preparation, logistics, enrollment, sampling, and shipping of samples for testing. We must also acknowledge dedication and commitment of US Navy Sailors and their respective commands without whom this project would not have been possible.

## Additional Information

### Disclaimers

All authors declare no competing or conflicts of interests. The views expressed in this article are those of the authors and should not be construed to reflect the official policy or represent the positions of the Department of the Navy, Department of the Army, Department of Defense, nor the United States Government. Material has been reviewed by the Walter Reed Army Institute of Research. There is no objection to its presentation and/or publication. The investigators have adhered to the policies for protection of human subjects as described in AR 70-25. LT Huy C. Nguyen, MC, USN NAMRU-IP; LCDR Robert D. Hontz, MSC, NAMRU-IP; LCDR Stephen E. Lizewski, MSC, NMRC; and CAPT Andrew G. Letizia, MC, USN NAMRU-IP are military service members. This work was prepared as part of my official duties. Title 17 U.S.C. 105 provides that ‘copyright protection under this title is not available for any work of the United States Government.’ Title 17 U.S.C. 101 defines a U.S. Government work as work prepared by a military service member or employee of the U.S. Government as part of that person’s official duties.

### Funding

This work was supported by funds from the US Bureau of Medicine and Surgery (BUMED) Restore Core Basic Operational Med Research Sciences to HCN and AGL. The study number is NV.6.R22.

### Contributions

Conceptualization: HCN, AGL, RDH; Draft Writing-original: all authors; Sample Collection: HCN, AGL, and Vysnova study team members; laboratory work: HSF, SEL, PS, CAB, KGL, SJK, LP, YC, IF, LS; Data analysis: PS, CAB, KGL, SJK; Data interpretation: HCN, AGL, PS, CAB, KGL, SJK; Writing-review and editing: all authors; Coordination: HCN and AGL.

## SUPPLEMENTAL FIGURES

**Figure S2.**
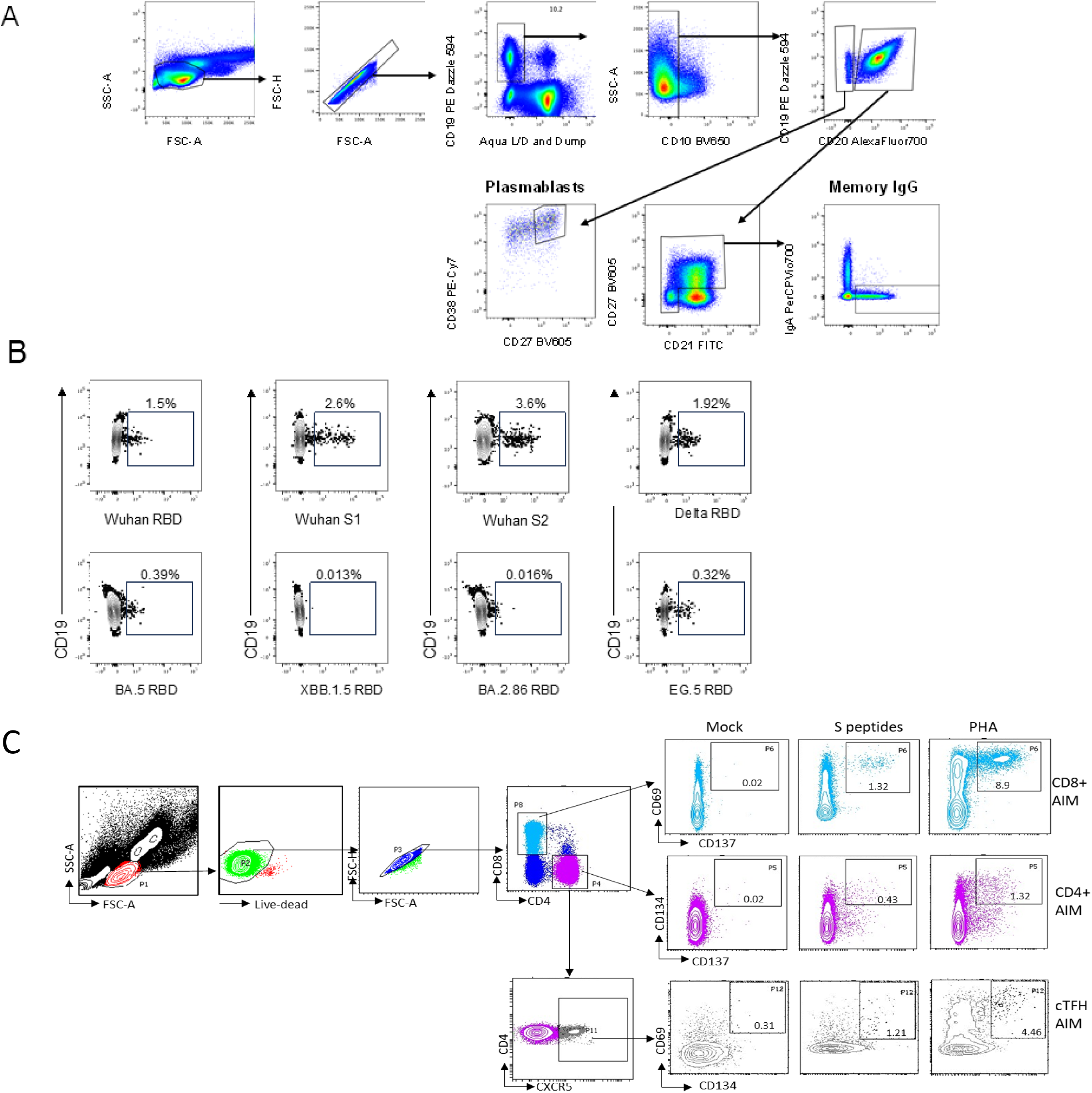
Gating strategy for AIM positive PBMCs. **A**, Example of gating strategy using flow cytometry for the identification of SARS-CoV-2-positive B cells within the memory IgG and plasmablast populations. Plasmablast populations were defined as CD19+, dump channel/viability dye negative, CD10-, CD20- and CD38bright CD27+ whereas memory populations were defined as CD19+, dump channel negative, CD10-, CD20+, CD27-/+ CD21/CD27 double positive. **B**, Example of gating strategy using flow cytometry for the identification of SARS-CoV-2-positive B cells within the memory IgG population in a LINKS donor. Gates were determined based on staining from a pre-pandemic, SARS-CoV-2 inexperienced donor. **C**, Gating strategy to quantify AIM positive T cells following antigen stimulation from a representative donor. CD8^+^ T cell populations were defined as CD69+CD137+ whereas CD4^+^ populations defined as CD134+CD137+.

**Figure S3.**
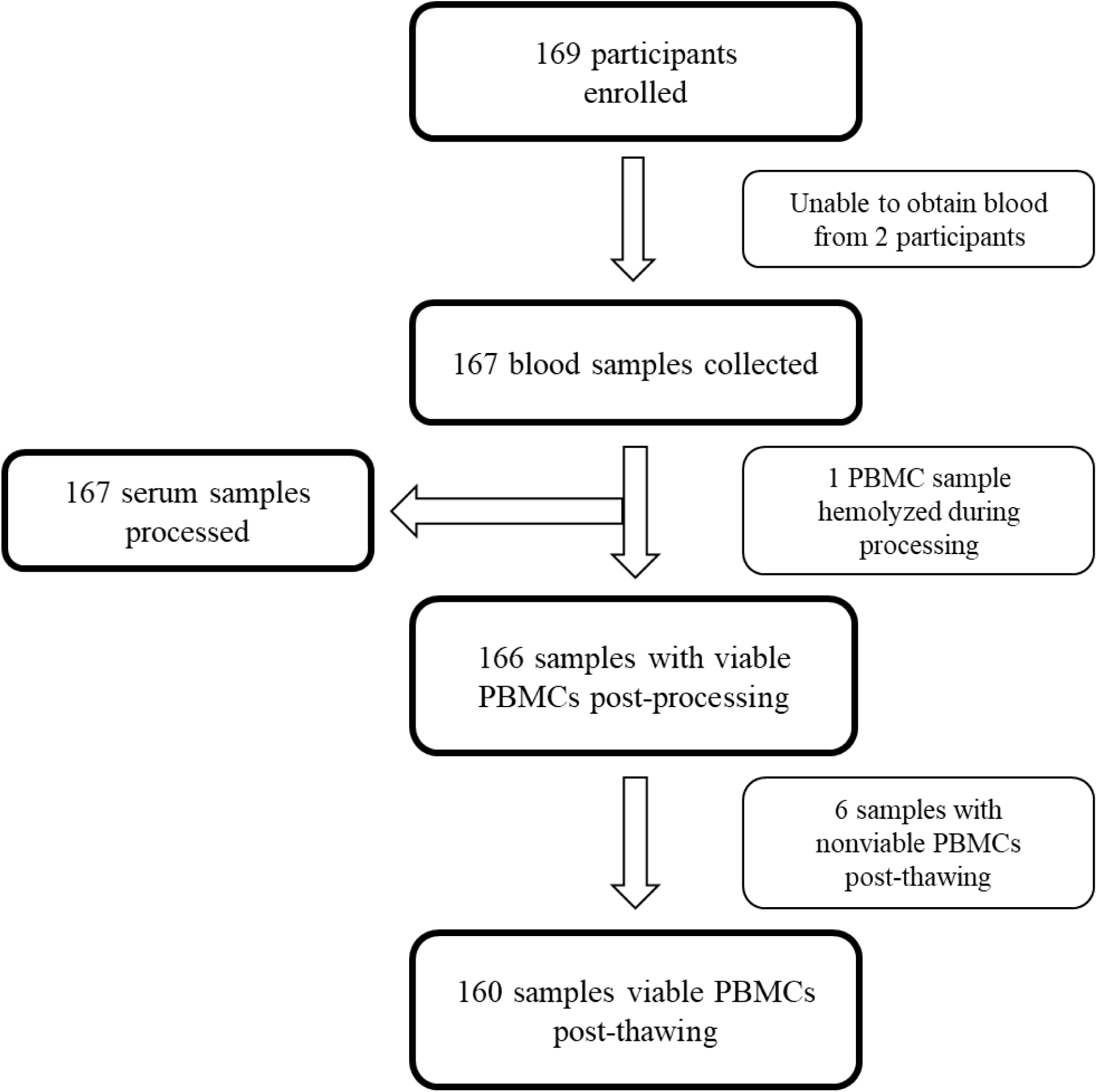
Number of enrolled participants and viable samples acquired.

**Figure S4.**
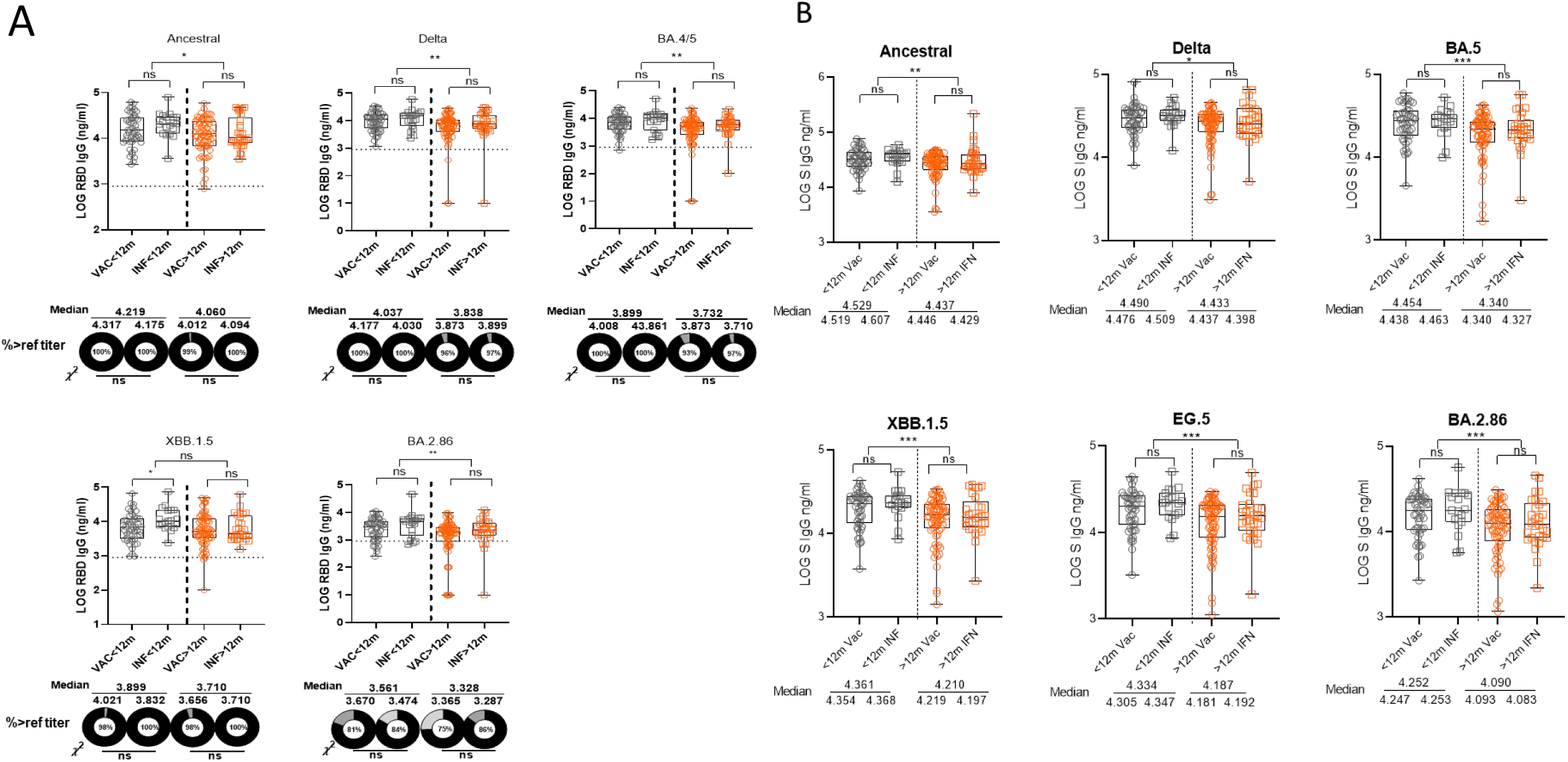
IgG titers to RBD and spike (S) protein of PDVs and CVs based on time (12-month cutoff) and nature of most recent exposure (either vaccine or infection) **A**, RBD IgG titers. The percentages of participants with titers above the reference “protective” titer (861 ng/ml or 2.94 in log scale) represented as dashed line are shown as donut plots. **B**, S IgG titers. 167 donors with sera. Black circles and squares represent participants with an exposure within 12 months of enrollment while orange circles and squares the participants with an exposure greater than 12 months from enrollment. Circles represent those whose most recent exposure was a vaccine while squares those whose most recent exposure was an infection. Median titers are shown below each group in ng/ml. Mann-Whitney U test was performed to show group median differences. Chi square analysis was performed between selected groups. Significant differences are marked as *p < 0.05, **p < 0.01, ***p < 0.001. Abbreviations: IgG, immunoglobulin G; RBD, receptor-binding domain; PDVs, Previous Dominant Variants (Ancestral, Delta, and BA.4/BA.5); CVs, Circulating Variants (XBB.1.5, EG.5, and BA.2.86).

**Figure S5.**
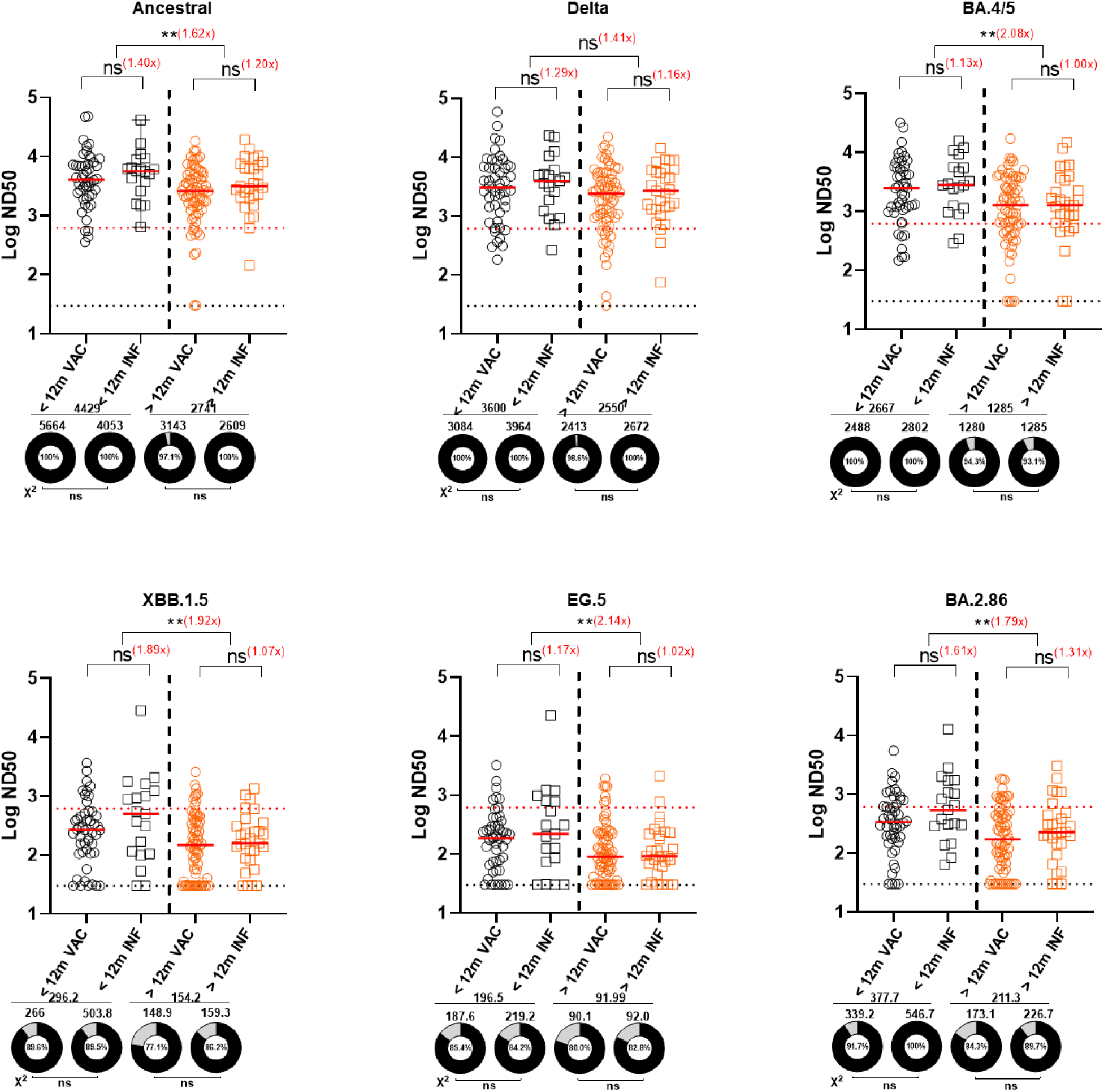
Pseudovirus neutralization antibody (NAb) titers to RBD of PDVs and CVs based on time (12-month cutoff) and nature of most recent exposure (either vaccine or infection) 167 donors with sera. Grey circles and squares represent participants with an exposure within 12 months of enrollment while orange circles and squares the participants with an exposure greater than 12 months from enrollment. Circles represent those whose most recent exposure was a vaccine while squares those whose most recent exposure was an infection. Log ND50 values are shown. Numbers next to group comparisons represent fold-change in median values. The percentages of participants with titers above the LOD (30 or 1.5 in log scale) represented as dashed line are shown as donut plots. Median titers are shown below each group in ng/ml. Mann-Whitney U test was performed to show group median differences. Chi square analysis was performed between selected groups. Significant differences are marked as *p < 0.05, **p < 0.01, ***p < 0.001. Abbreviations: RBD, Receptor-Binding Domain; PDVs, Previous Dominant Variants (Ancestral, Delta, and BA.4/BA.5); CVs, Circulating Variants (XBB.1.5, EG.5, and BA.2.86).

**Figure S6.**
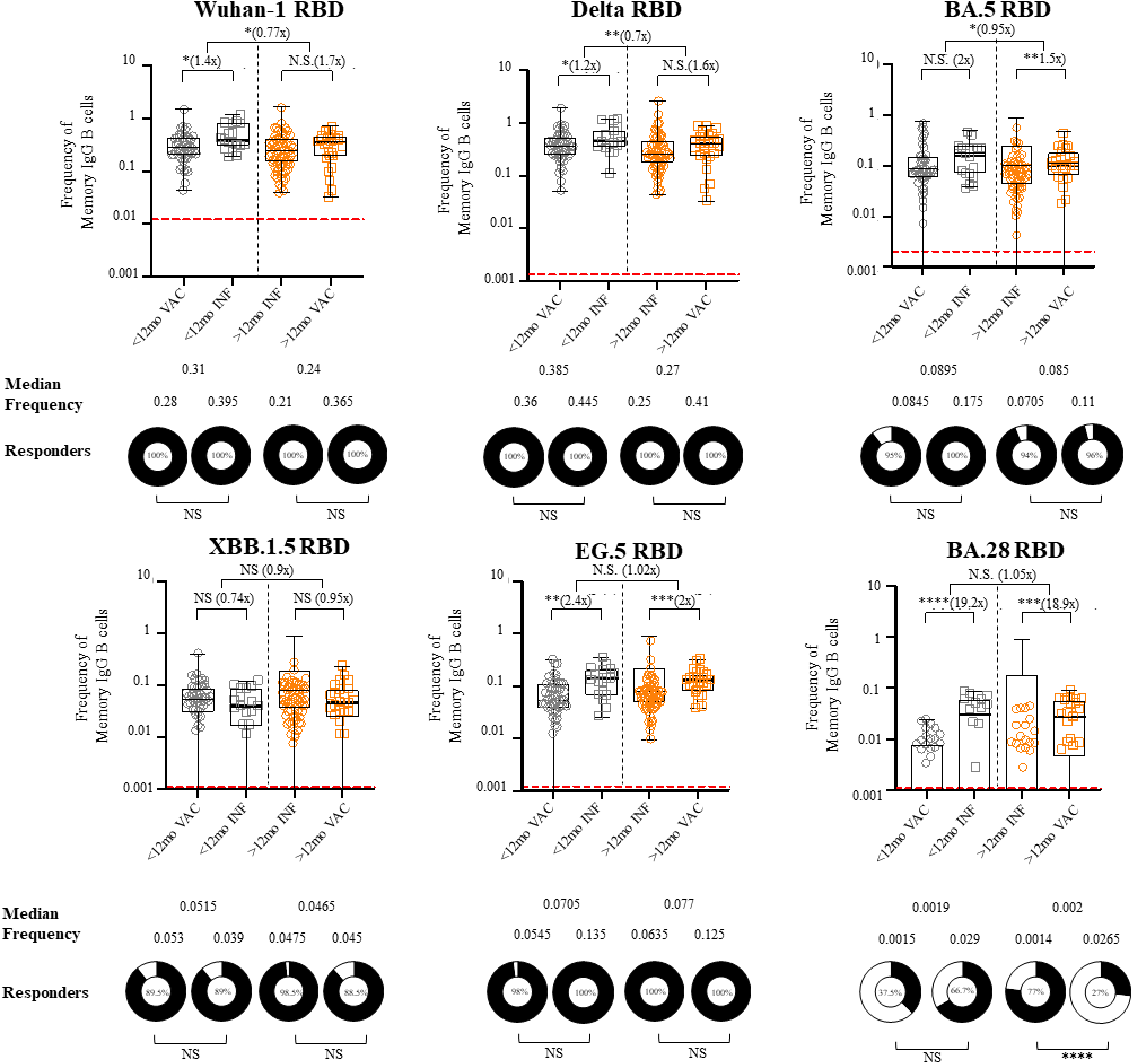
Frequency of IgG positive memory B cells (MBCs) to RBD of PDVs and CVs based on time (12-month cutoff) and nature of most recent exposure (either vaccine or infection) 160 donors with PBMCs. Grey circles and squares represent participants with an exposure within 12 months of enrollment while orange circles and squares the participants with an exposure greater than 12 months from enrollment. Circles represent those whose most recent exposure was a vaccine while squares those whose most recent exposure was an infection. Numbers next to group comparisons represent fold-change in median values. The percentages of MBC “responders” are shown as donut plots. MBC “responder” is defined as having a frequency higher than the negative control sample run on the same day. The red dashed line represents the median of the frequencies of MBCs meeting gating criteria per variant across all days of analysis from a negative control donor. Median frequencies are shown below each group. Mann-Whitney U test was performed to show group median differences. Chi square analysis was performed between selected groups. Significant differences are marked as *p < 0.05, **p < 0.01, ***p < 0.001, ****p<0.0001. Abbreviations: IgG, immunoglobulin G, RBD, Receptor-Binding Domain; PDVs, Previous Dominant Variants (Ancestral, Delta, and BA.4/BA.5); CVs, Circulating Variants (XBB.1.5, EG.5, and BA.2.86); MBC, memory B cells; PBMCs, peripheral blood mononuclear cells.

**Figure S7.**
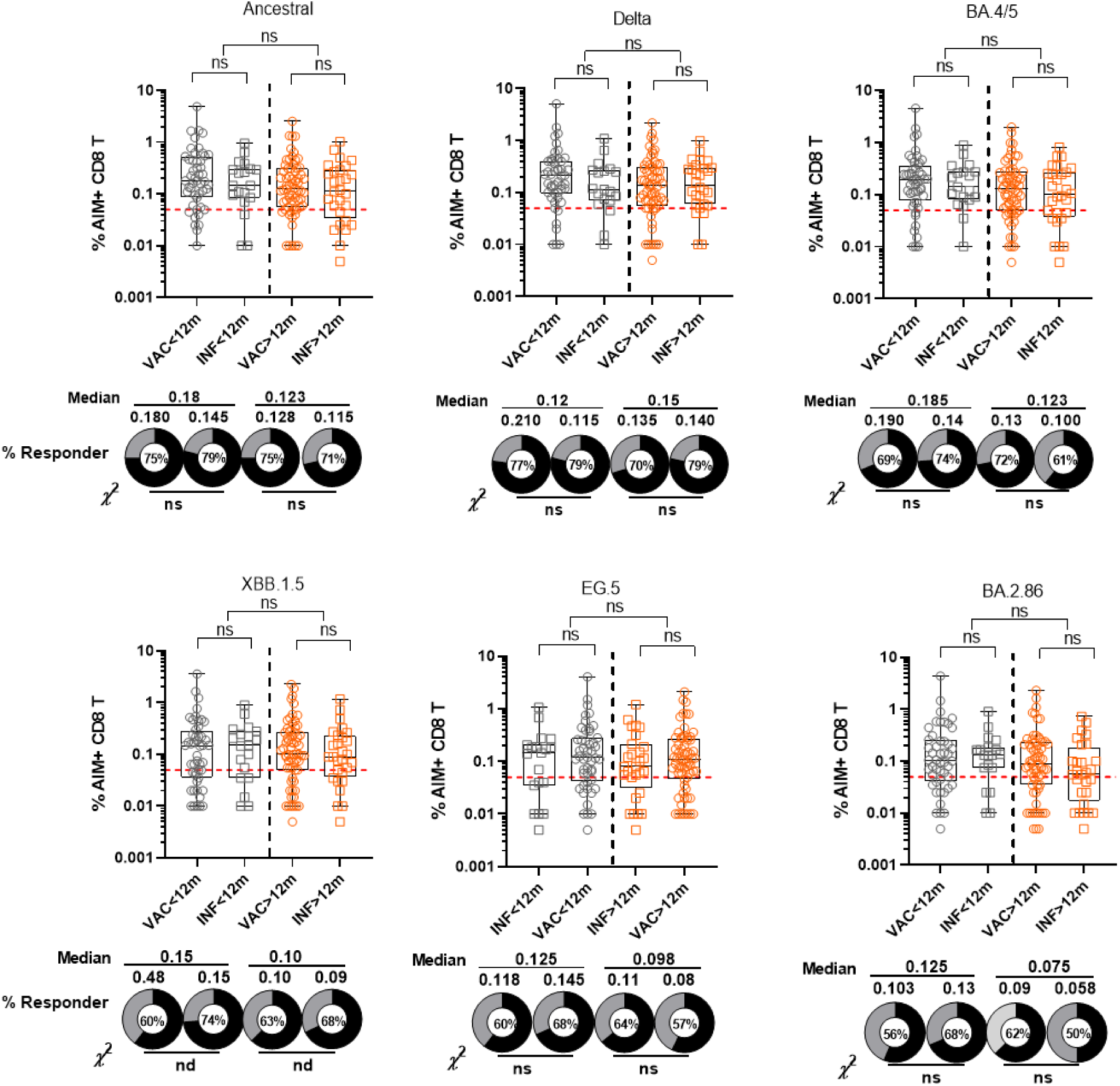
Frequencies of AIM positive CD8+ T cells to RBD of PDVs and CVs based on time (12-month cutoff) and nature of most recent exposure (either vaccine or infection) 160 donors with PBMCs. Grey circles and squares represent participants with an exposure within 12 months of enrollment while orange circles and squares the participants with an exposure greater than 12 months from enrollment. Circles represent those whose most recent exposure was a vaccine while squares those whose most recent exposure was an infection. AIM positivity defined as CD69+ and CD137+. The percentages of CD8+ T cell “responders” are shown as donut plots. CD8+ T cell “responder” is defined as having a frequency higher than 0.05% and a stimulation index of greater than 2. Median frequencies are shown below each group. Mann Whitney U test was performed to show group median differences. Chi square analysis was performed between selected groups. Significant differences are marked as *p < 0.05, **p < 0.01, ***p < 0.001. Abbreviations: IgG, immunoglobulin G, RBD, Receptor-Binding Domain; PDVs, Previous Dominant Variants (Ancestral, Delta, and BA.4/BA.5); CVs, Circulating Variants (XBB.1.5, EG.5, and BA.2.86); MBC, memory B cells; PBMCs, peripheral blood mononuclear cells.

**Figure S8.**
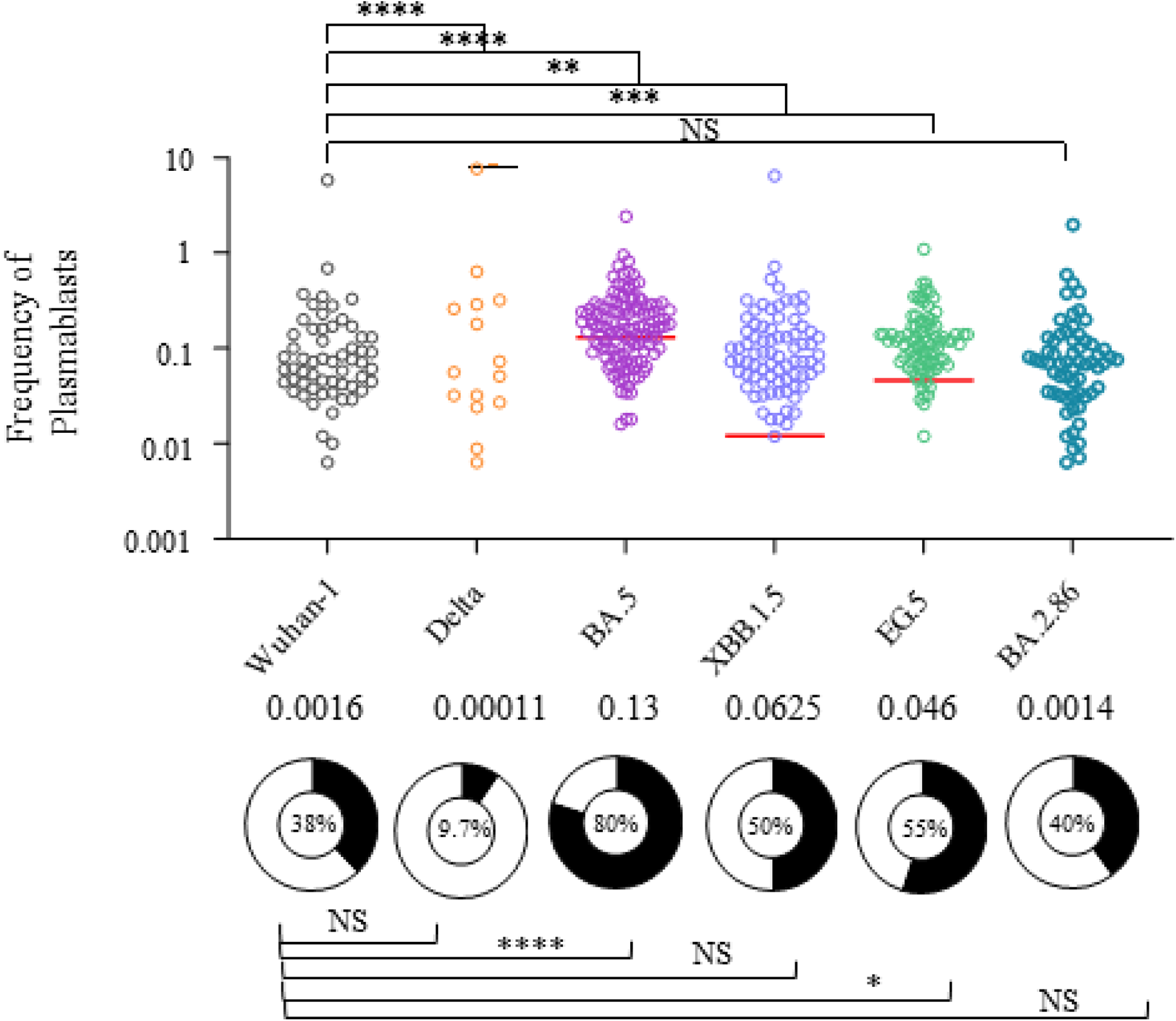
Frequency of plasmablasts to RBD of PDVs and CVs. 160 donors with PBMCs. The percentages of plasmablasts “responders” are shown as donut plots. Plasmablast “responder” is defined as having a plasmablast frequency higher than the negative control sample run on the same day. The dashed line represents the median of the frequencies of plasmablasts meeting gating criteria per variant across all days of analysis from a negative control donor. Median frequencies are shown below each group. Wilcoxon signed-rank test was performed to show group median differences. Significant differences are marked as *p < 0.05, **p < 0.01, ***p < 0.001, ****p<0.0001. Abbreviations: IgG, immunoglobulin G, RBD, Receptor-Binding Domain; PDVs, Previous Dominant Variants (Ancestral, Delta, and BA.4/BA.5); CVs, Circulating Variants (XBB.1.5, EG.5, and BA.2.86; PBMCs, peripheral blood mononuclear cells.

**Figure S9.**
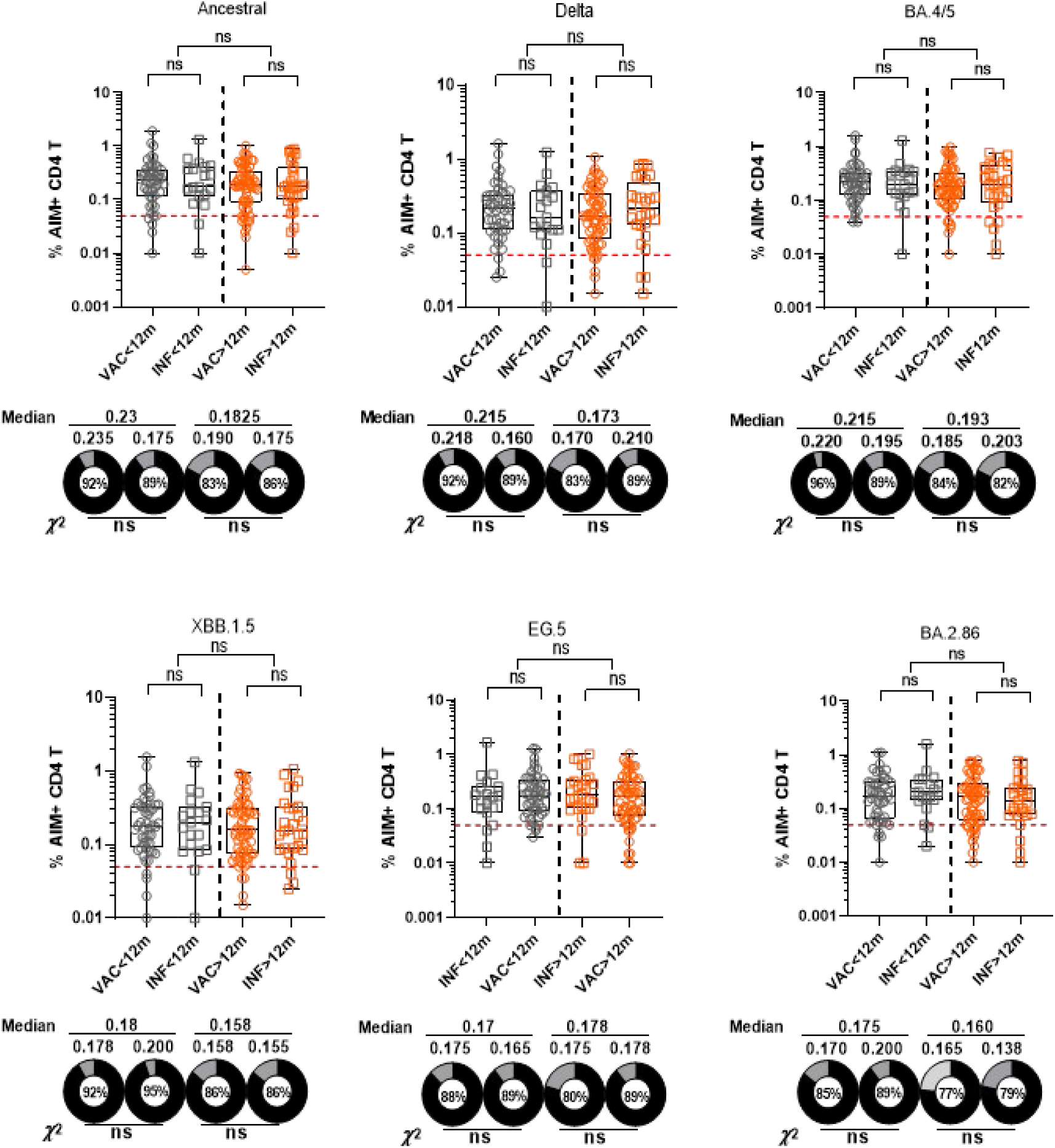
Frequencies of AIM positive CD4+ T cells to RBD of PDVs and CVs based on time (12-month cutoff) and nature of most recent exposure (either vaccine or infection) 160 donors with PBMCs. Grey circles and squares represent participants with an exposure within 12 months of enrollment while orange circles and squares the participants with an exposure greater than 12 months from enrollment. Circles represent those whose most recent exposure was a vaccine while squares those whose most recent exposure was an infection. AIM positivity defined as CD134+ and CD137+. The percentages of CD4+ T cell “responders” are shown as donut plots. CD4+ T cell “responder” is defined as having a frequency higher than 0.05% and a stimulation index of greater than 2. Median frequencies are shown below each group. Mann Whitney U test was performed to show group median differences. Chi square analysis was performed between selected groups. Significant differences are marked as *p < 0.05, **p < 0.01, ***p < 0.001. Abbreviations: IgG, immunoglobulin G, RBD, Receptor-Binding Domain; PDVs, Previous Dominant Variants (Ancestral, Delta, and BA.4/BA.5); CVs, Circulating Variants (XBB.1.5, EG.5, and BA.2.86); MBC, memory B cells; PBMCs, peripheral blood mononuclear cells.

## Notes

### Competing Interest Statement

The authors have declared no competing interest.

### Author Declarations

IRB of US Naval Medical Research Command (Silver Spring, MD, USA) gave ethical approval for this work.

