## Supplemental Figure 1 and Supplemental Methods for "Informing the need for a SARS-CoV-2 booster based upon the immune response among young, healthy adults to variants circulating during late 2023"

**Figure S1. Study questionnaire regarding demographics, COVID-19 exposure history, and vaccine attitudes of participants.**

| | |
|---|---|
| Date of enrollment:<br>☐☐/☐☐/☐☐☐☐ (MM/DD/YYYY) | |
| Date of sample collected:<br>☐☐/☐☐/☐☐☐☐ (MM/DD/YYYY) | Date arrived in USINDOPACOM<br>☐☐/☐☐/☐☐☐☐ (MM/DD/YYYY) |

| **A. DEMOGRAPHIC**<br>**Please select only one correct answer by checking on the tick box** | | |
|---|---|---|
| **Code** | **Question** | **Answer** |
| A1 | DOD ID | |
| A2 | Date of birth | ☐☐/☐☐/☐☐☐☐ (MM/DD/YYYY) |
| A3 | Gender | ☐1 Male ☐2 Female |
| A4 | Ethnicity | ☐1 Hispanic or Latino ☐2 Not Hispanic or Latino |
| A5 | Race | ☐1 American Indian or Alaska Native ☐2 Asian<br>☐3 Black/African American ☐4 Native Hawaiian/Pacific Islander<br>☐5 White |
| A6 | Weight | ☐☐☐ pounds A7 Height ☐ feet, ☐☐ inches |
| A8 | Service | ☐1 Army ☐2 Marines ☐3 Navy<br>☐4 Air Force ☐5 Other: ________________ |
| A9 | Rank/grade | ☐1 E1 – E4 ☐2 E5 – E6 ☐3 E7 – E9<br>☐4 Warrant ☐5 O1 – O3 ☐6 O4 – O6 |
| A10 | Military specialty: | |

| **B. COVID-19 VACCINATION**<br>**Please answer all questions** | | |
|---|---|---|
| **Code** | **Question** | **Answer** |
| B1 | Which COVID-19 vaccine **primary series** did you receive?<br>DO NOT COUNT boosters | ☐1 Pfizer/BioNTech BNT162b2 – 2 doses<br>☐2 Moderna mRNA-1273 – 2 doses<br>☐3 Novavax NVX-CoV2373 – 2 doses<br>☐4 Johnson & Johnson Ad26.COV2.S – 1 dose<br>☐5 A combination of the above – please specify brand and dose: |
| B2 | Date of **completion** of primary series (provide BEST estimate in DDMMMYYYY; please utilize *proofs of record* such as vaccine cards if available) | |
| B3 | Have you received a COVID-19 **booster** vaccine (if answer is No, skip to C1)? | ☐1 Yes ☐2 No |
| B4 | **Name and number** of vaccine booster ever received (select all applicable options) | ☐1 Pfizer/BioNTech – 1 booster dose<br>☐2 Pfizer/BioNTech – 2 booster doses<br>☐3 Pfizer/BioNTech – 3 booster doses<br>☐4 Moderna – 1 booster dose<br>☐5 Moderna – 2 booster doses<br>☐6 Moderna – 3 booster doses |
| B5 | **TOTAL** number of booster doses received | |
| B6 | Have you received the new **BIVALENT** COVID-19 vaccine booster (either Pfizer/BioNTech or Moderna)? (This booster targets both the original SARS-CoV-2 strain and the **Omicron B.4/B.5** variants, approved by FDA August 2022) | ☐1 Yes ☐2 No |
| B7 | **Type and date** of receipt of booster vaccines (for type, select either Monovalent or Bivalent; for date, provide BEST estimate in MMDDYYYY; please utilize *proofs of record* such as vaccine cards if available) | **BOOSTER #1**:<br>☐1 Monovalent<br>☐2 Bivalent<br>☐3 Date:<br><br>**BOOSTER #2**:<br>☐1 Monovalent<br>☐2 Bivalent<br>☐3 Date:<br><br>**BOOSTER #3**:<br>☐1 Monovalent<br>☐2 Bivalent<br>☐3 Date: |

| C. PRIOR COVID-19 INFECTION<br>Please answer all questions | | |
|---|---|---|
| **Code** | **Question** | **Answer** |
| C1 | Have you ever had a **positive** COVID-19 test (if answer is No, skip to D1)? | ☐1 Yes ☐2 No |
| C2 | How many **separate** times have you been infected with COVID-19 either with or without symptoms? (Positive tests separated by more than 90 days WITH normal state of health in between episodes) | |
| C3 | Which COVID test platform was used for the positive test result(s) (select all applicable options)? | ☐1 Antigen-based (rapid or home test kits) – nose swab<br>☐2 PCR-based – nose swab<br>☐3 Antibody-based - blood draw |
| C4 | Date(s) of positive COVID-19 test(s) (provide best estimate in DDMMMYYYY) | **Episode #1**:<br><br>**Episode #2** (if applicable):<br><br>**Episode #3** (if applicable): |
| C5 | Did you seek medical attention during any COVID-related episodes? | ☐1 Yes ☐2 No |
| C6 | Did you ever have to miss work due to any COVID-related episodes (if answer is No, skip the next question)? | ☐1 Yes ☐2 No |
| C7 | How many days did you have to miss work in total? | |
| C8 | Were you ever hospitalized during any COVID-related episodes? | ☐1 Yes ☐2 No |

| D. HEALTH RISKS<br>Please answer all questions by checking on the tick box | | | |
|---|---|---|---|
| **Code** | **Question** | **Answer** | **Guide** |
| D1 | Do you currently use tobacco products? | ☐1 Yes ☐2 No | **If No, skip to D2** |
| *D1a* | Cigarettes - # Cigarettes per day (average) | ☐☐☐ (cigarettes) | |
| *D1b* | Vaping (e-cigarettes) - # e-cig use sessions per day (assume that 1 session consists of 15 puffs OR lasts around 10 minutes) | ☐☐☐ (sessions) | |
| *D1c* | Chewing - # Pouches per day (average) | ☐☐☐ (pouches) | |
| *D1d* | Dipping - # Cans of snuff per day (average) | ☐☐☐ (cans) | |
| *D1e* | How long have you been using tobacco products? | ☐☐ ☐ (months) | |
| D2 | Have you ever used tobacco products in the past? | ☐1 Yes ☐2 No | **If No, skip to D3** |
| D2a | How long did you use tobacco products for? | ☐☐ ☐ (months) | |
| D2b | Which tobacco product(s) did you use? Check all that apply. | ☐1 Cigarettes<br>☐2 Smokeless tobacco (chewing, dipping)<br>☐3 Vaping (e-cigarettes) | |
| D2c | When did you stop using tobacco product? (DDMMMYYYY) | | |
| D3 | Take medicine daily | ☐1 Yes ☐2 No | |
| *D3a* | What type of medicines/drugs you are taking? | | |

| E. ATTITUDES regarding VACCINATION<br>Please answer all questions | | |
|---|---|---|
| **Code** | **Question** | **Answer** |
| E1 | If there is a new FDA-approved booster vaccine against the most commonly circulating COVID-19 variant, how likely are you to obtain it if it is not mandated by the DoD? | ☐1 Highly unlikely<br>☐2 Not very likely<br>☐3 Neutral<br>☐4 More likely than not<br>☐5 Highly likely |
| E2 | If your answer above is “highly unlikely,” “not very likely,” or “neutral,” what are your reasons and/or concerns? Check all that apply. | ☐1 Adverse side effects from the boosters<br>☐2 Anxiety/fear of needles<br>☐3 Boosters are not very effective<br>☐4 Boosters do not provide additional benefit beyond primary series<br>☐5 COVID-19 is no longer a major concern<br>☐6 Other (please specify): |
| E3 | Out of the above reasons/concerns, which is MOST important to you? | |
| E4 | If the new booster can be administered **orally** (via a pill or tablet) or **intranasally** (via inhalation device, drops, or spray) without using any sharps, how likely are you to obtain it if it not mandated? (Assume this booster is FDA-approved) | ☐1 Highly unlikely<br>☐2 Not very likely<br>☐3 Neutral<br>☐4 More likely than not<br>☐5 Highly likely |
| E5 | For future DoD-mandate or DoD-recommended vaccines, which route of administration do you prefer? (Assume all vaccines are FDA-approved) | ☐1 Intramuscular/subcutaneous via needles<br>☐2 Oral (pill/tablet)<br>☐3 Intranasal (inhaled, drops, spray) |
| E6 | Does the annual influenza (flu) vaccine provide protection against COVID-19? | ☐1 Yes ☐2 No |

### Supplemental Methods

##### Study Design, Enrollment, and Sampling

Thirty-five milliliters (35 mL) of whole blood were collected from each study participant via venipuncture and processed on site within 4 hours and frozen at -80°C. All samples were shipped to two DoD laboratories (Naval Medical Research Command and Walter Reed Army Institute of Research) in Silver Spring, Maryland within 3 weeks from the first enrollment event.

##### Quantitative ELISA of Receptor-Binding Domain (RBD) and Spike (S) Protein IgG

Antigens used for ELISA were all from ACRO Biosystems. Trimeric S proteins and RBD from Ancestral (SPN-C52H9), Delta (SPN-C52He), BA.4/5 (SPN-C522e), XBB.1.5 (SPN-C524i), EG.5 (SPN-C524u), and BA.2.86 (SPN-C524y), RBD from Ancestral (SPD-C52H3), Delta (SPD-C52Hh), BA.4/5 (SPD-C522r), XBB.1.5 (SPN-C5242), and BA.2.86 (SPD-C5247) were used. All antigens were diluted in 1x phosphate buffered saline without Ca++ and Mg++ (1xdPBS) and ELISA plates were coated with 1ug/ml (100 ul) of antigens overnight. ELISA was performed as previously described [13]. Plates coated with dPBS were used to assess non-antigen-specific binding. Prior to immunoassay, serum samples were heat-inactivated at 56°C for 1 h. Samples were diluted in dPBS and 100 ul of samples were added to the ELISA plates. For S and RBD, we measured IgA and IgG. For IgA and IgG standard curves, microplates were coated with 1 μg/ml (0.1 ug/well) goat anti-human IgA (Southern Biotech; Cat: 2050-01) and IgG (Southern Biotech; Cat: 2040-01) in dPBS at 4˚C overnight. After washing in 1×dPBS-Tween-20, plates were blocked with 5% skim milk in dPBS at 37˚C for 1 h. Purified IgA (Athens Research; Cat#: 16-16-090701), and IgG (Athens Research; Cat#: 16-16-090707) were 2-fold serially diluted with Sample Dilution Buffer as standard curves (ranging from 0.4 ng/ml to 800 ng/ml). All plates were then incubated at 37˚C for 2 h.

After washing, the plates were incubated with 1:1,000 diluted goat anti-human IgA-HRP (Southern Biotech; Cat: 2050-05) or IgG-HRP (Southern Biotech; Cat: 2040-05), at 37˚C for 1 h. HRP was detected with TMB Microwell Peroxidase substrate (SeraCare; Cat: 5120-0077) and the reactions were stopped by 1N sulfuric acid (H2SO4) solution (Fisher, Cat: SA212-1). Plates were read at OD450nm on plate reader (PerkinElmer EnSpire) within 30 min of stopping. We subtracted responses against the non-specific binding (PBS-coated wells) during data finalization to yield antigen-specific response. SARS-CoV-2-specific IgA, IgM or IgG concentrations (ng/ml) were calculated by sigmoidal standard curves using GraphPad Prism 8 and converted to ng/ml based on the dilution factor.

For RBD IgG, a cutoff of 861 ng/ml (corresponding to 2.94 on the log scale) was used as a "responder" threshold, represented as a dashed line in Figure 1. This value was calculated as an average of two RBD IgG titers (816 ng/ml and 906 ng/ml) previously reported in the literature to correlate with high vaccine efficacy rate and clinically significant host viral neutralizing capacity; thus, they could be considered as surrogate markers of protection [27,28].

##### Pseudovirus Neutralization Assay

Pseudovirus (PV) was produced using the SARS-CoV-2 Spike (S) gene and the lentivirus derived reporter and packaging plasmids previously described [29], and generously made available through BEI resources (NIAID, NIH). Spike gene sequences were obtained from the GSAID database (www.gsaid.org), codon optimized for human expression with 19 amino acids removed from the C-terminus (GeneArt, ThermoFisher), and cloned into a pcDNA3.1+ expression vector. PV neutralization assays were performed as previously described [30]. Serum was heat inactivated at 56°C for one hour prior and sample serum diluted 1:30 (set as limit of detection) in DMEM containing 10% FBS, followed by three-fold serial dilutions in black, flat bottom 96-well plates (Greiner BIO-ONE). After 48-hour incubation, luciferase activities were measured using Pierce Firefly Luc One-Step Glow Assay Kit, according to the manufacturer's instructions. Luminescence was read using an

EnSpire plate reader (PerkinElmer). Data was analyzed using Prism GraphPad software (Version 8.3.1), using nonlinear regression analysis to calculate ND50. The limit of detection of the assay was 30.

**Memory B Cells Frequencies and Responses**

Cryopreserved PBMCs from the LINKS study were thawed in warm media containing benzonase, then washed with PBS and stained for viability using the Aqua Live/Dead stain (ThermoFisher) and Fc receptors were blocked using normal mouse IgG (Invitrogen #02-6502). After centrifuging and removal of supernatant, cells were stained with fluorochrome conjugated secondary antibodies specific to subdomains of the S protein of SARS-CoV-2 using fluorochrome conjugated streptavidin tetramers conjugated to the biotinylated RBDs of Wuhan 1 (Acrobiosystems cat# SPD-C82E9-25ug), Delta B.1.617.2 (Acrobiosystems cat# SPD-C82Ed-25ug), Omicron BA.5 (Acrobiosystems cat# SPD-C82Ew-25ug), XBB.1.5 (Acrobiosystems cat# SPD-C82Q3-25ug), BA.2.86 Acrobiosystems cat# SPD-C5247-100ug), EG.5 (Acrobiosystems cat# SPD-C82Q5-200ug), and full-length Wuhan S1 (SinoBiological cat# 40591-V08H) and S2 (Sinobiologicals cat# 40150-V08B3), together with a cocktail of phenotyping antibodies including CD20 AlexaFluor700 (BD Biosciences cat# 560631), IgD APC-H7 (BD Biosciences cat# 561305), IgM BUV395 (BD Biosciences cat# 563903), CD4 BV510 (BD Biosciences cat# 562970), CD16 BV510 (BD Biosciences cat# 563830), CD10 BV650 (BD Biosciences cat# 563734), IgG BUV495 (BD Biosciences cat#741172), CD14 BV510 (BioLegend cat#301842), CD3 BV510 (BioLegend cat#300448), CD56 BV510 (BioLegend cat#362534), CD8a BV510 (BioLegend cat#301048), CD27 BV605 (BioLegend cat#302830), CD21 FITC (BioLegend cat#354910), CD38 PECy7 (BioLegend cat# 303516), CD19 PEDazzle 594 (BioLegend cat# 302252). SARS-CoV-2 antigen positive B cells were identified amongst plasmablast populations, defined as CD19+, dump channel/viability dye negative, CD10-, CD20- and CD38$^{bright,}$ CD27+, and memory IgG populations defined as CD19+, dump channel negative, CD10-, CD20+, CD27+ CD21-/+. Responders within each SARS-CoV-2 antigen tetramer were identified as individuals with positive frequencies within either the memory IgG or plasmablast gate above that of a negative control (pre-pandemic COVID-19) donor run on the same assay day with LINKS sample sets to draw gates.

**T Cells Frequencies and Responses**

PBMCs were thawed and counted for cell viability. Among 167 study subjects, samples from 7 subjects yielded viability <40%, and were excluded from further usage. Samples from the rest of 159 subjects yielded an average viability of 92.6% with a standard deviation (SD) of 18.9 and were used for setting up the AIM assays on the same day.

Assay methods for AIM were described previously [31]. Briefly, PBMCs were cultured in the presence of SARS-CoV-2-specific S peptide pools (1mg/mL) in 96-well U-bottom plates at a concentration of 1ug/ml per 500,000 PBMCs in duplicate wells. The S protein peptide pools used in the study were all from JPT Peptide Technologies GmbH and included Ancestral (Cat#: PM-WCPV-S1), Delta (Cat#:PM-SARS2-SMUT06-1), BA.4/5 (Cat#: PM-SARS-2-SMUT10-1), XBB.1.5 (Cat#: PM-SARS2-SMUT15-1), EG.5 (Cat#: PM-SARS-2-SMTU16-1), and BA.2.86 (Cat#: PM-SARS2_SMUT17-1). The negative control was an equimolar of DMSO. For a positive control, phytohemagglutinin (PHA, Roche,5ug/mL) was used to stimulate cells. We have included a pre-pandemic sample from a dengue-exposed subject and stimulated this sample with the same SARS-CoV-2 peptide pools and a Dengue peptide pool during the setup of each assay. If negative responses to SARS-CoV-2 peptide pools and positive responses to the dengue peptide pool were observed, the assay was accepted.

After antigen stimulation for 15-18 h at 37 ºC in 5% CO2, PBMCs were washed and stained for 30 min on ice with a cocktail antibodies: CD4-APC-H7 (clone SK3, BD; Cat# 641398), CD8-PerCP (clone SK1, BD Cat#347314), CD134-PE-Cy7 (clone ACT35, Bio Legend; Cat #:350012), CD137-APC (BD; Cat#: 550890), CD69-BV421 (BD; Cat#562884), CD25-PE (BD; Cat#555432), CXCR5-AlexaFluor-488 (clone RF8B2, BD; Cat # 558112), and a live-dead dye (aqua fluorescent reactive dye for 405nM excitation, Invitrogen; L34966A) for CD4 and CD8 T cells. After staining, cells were then acquired directly on a FACS CANTO II with 3-lasers (BD) and analyzed with FlowJo software (Tree Star Inc.).

The gating strategy for AIM cells was drawn relative to the negative and positive controls for each donor as shown in Supplementary Figure S2. Lymphocytes were first gated on FCS-A vs SSC-A and dead cells were excluded by gating on Live-dead dye. Single cells were selected by excluding doublets using FSC-A and FSC-H. Next, CD4+ and CD8+ T cells were gated. Activated CD4 T cells were further gated on CD134+CD137+ (CD4 AIM) or CD134+CD25+ (a broader AIM); activated CD8 T cells were gated on CD69+CD137+ AIM markers. The antigen activated CD4 and CD8 cells were presented as % CD134+CD137+/total CD4 cells and %CD137+CD69+/total CD8 cells, respectively.

The percentages of the AIM+ cells in duplicate wells were averaged. The antigen-specific AIM responses were obtained by subtracting the AIM responses in DMSO cultures. The Stimulation Index (SI) was calculated by dividing the % of AIM + cells in peptides-stimulated cultures against the % of AIM+ cells in the DMSO control. To obtain the cut-off for positive response, we used 6 pre-pandemic PBMC samples and stimulated the samples with the same antigens. After subtracting the responses in DMSO, we yielded antigen-stimulated responses from these 6 pre-pandemic subjects. We then obtained the mean and SD from all the responses and calculated the cut-off as mean+1.7 SD [32]. The cut-offs for both CD4 AIM and CD8 AIM were 0.05%. A response was considered positive when the SI was greater than 2 and the value after subtracting DMSO was above the cut-off value.

| Reagent | Source | Identifier |
|---|---|---|
| **Chemicals, proteins, and antigens** | | |
| TMB Microwell Peroxidase substrate | SeraCare | Cat# 5120-0077 |
| 1N sulfuric acid (H2SO4) solution | Fisher Scientific | Cat# SA212-1 |
| LIVE/DEAD™ Fixable Aqua Dead Cell Stain Kit | Invitrogen (Thermo Fisher Scientific) | Cat# L34966 |
| Immunoglobulin M (IgM) | Athens Research & Technology | Cat# 16-16-090713-M |
| Immunoglobulin A (IgA) | Athens Research & Technology | Cat# 16-16-090701 |
| Immunoglobulin G (IgG) | Athens Research & Technology | Cat# 16-16-090707 |
| Mouse IgG | Invitrogen | Cat# 02-6502 |
| Antigens (ELISA) | | |
| Ancestral SARS-CoV-2 S Protein | ACROBiosystems | Cat# SPN-C52H9 |
| Delta (B.1.617.2) SARS-CoV-2 S Protein | ACROBiosystems | Cat# SPN-C52He |
| Omicron (BA.4/5) SARS-CoV-2 S Protein | ACROBiosystems | Cat# SPN-C522e |
| Omicron (XBB.1.5) SARS-CoV-2 S Protein | ACROBiosystems | Cat# SPN-C524i |
| Omicron (EG.5) SARS-CoV-2 S Protein | ACROBiosystems | Cat# SPN-C524u |
| Omicron (BA.2.86) SARS-CoV-2 S Protein | ACROBiosystems | Cat# SPN-C524y |
| Ancestral SARS-CoV-2 S Protein RBD | ACROBiosystems | Cat# SPD-C52H3 |
| Delta (B.1.617.2) SARS-CoV-2 S Protein RBD | ACROBiosystems | Cat# SPD-C52Hh |
| Omicron (BA.4/5) SARS-CoV-2 S Protein RBD | ACROBiosystems | Cat# SPD-C522r |
| Omicron (XBB.1.5) SARS-CoV-2 S Protein RBD | ACROBiosystems | Cat# SPD-C5242 |
| Omicron (BA.2.86) SARS-CoV-2 S Protein RBD | ACROBiosystems | Cat# SPD-C5247 |
| Omicron (B.1.1.529) SARS-CoV-2 S Protein Nucleocapsid protein | ACROBiosystems | Cat# NUN-C52Ht |
| Antigens (Memory B Cells) | | |
| Wuhan 1 SARS-CoV-2 S protein RBD | ACROBiosystems | Cat# SPD-C82E9-25ug |
| Delta (B.1.617.2) SARS-CoV-2 S protein RBD | ACROBiosystems | Cat# SPD-C82Ed-25ug |
| Omicron (BA.5) SARS-CoV-2 S protein RBD | ACROBiosystems | Cat# SPD-C82Ew-25ug |
| Omicron (XBB.1.5) SARS-CoV-2 S protein RBD | ACROBiosystems | Cat# SPD-C82Q3-25ug |
| Omicron (BA.2.86) SARS-CoV-2 S protein RBD | ACROBiosystems | Cat# SPD-C5247-100ug |
| Omicron (EG.5) SARS-CoV-2 S protein RBD | ACROBiosystems | Cat# SPD-C82Q5-200ug |
| Wuhan Spike S1 | SinoBiological | Cat# 40591-V08H |

| | | |
|---|---|---|
| Wuhan Spike S2 | SinoBiological | Cat# 40150-V08B3 |
| Antigens (T Cells) | | |
| Ancestral SARS-CoV-2 S Protein | JPT Peptide Technologies | Cat# PM-WCPV-S1 |
| Delta (B.1.617.2) SARS-CoV-2 S Protein | JPT Peptide Technologies | Cat# PM-SARS2-SMUT06-1 |
| Omicron (BA.4/5) SARS-CoV-2 S Protein | JPT Peptide Technologies | Cat# PM-SARS-2-SMUT10-1 |
| Omicron (XBB.1.5) SARS-CoV-2 S Protein | JPT Peptide Technologies | Cat# PM-SARS2-SMUT15-1 |
| Omicron (EG.5) SARS-CoV-2 S Protein | JPT Peptide Technologies | Cat# PM-SARS-2-SMTU16-1 |
| Omicron (BA.2.86) SARS-CoV-2 S Protein | JPT Peptide Technologies | Cat# PM-SARS2_SMUT17-1 |
| **Antibodies** | | |
| Goat anti-human IgM | SouthernBiotech | Cat# 2020-01 |
| Goat anti-human IgA | SouthernBiotech | Cat# 2050-01 |
| Goat anti-human IgG | SouthernBiotech | Cat# 2040-01 |
| Goat anti-human IgM-HRP | SouthernBiotech | Cat# 2020-05 |
| Goat anti-human IgA-HRP | SouthernBiotech | Cat# 2050-05 |
| Goat anti-human IgG-HRP | SouthernBiotech | Cat# 2040-05 |
| Mouse anti-human CD20 Alexa Fluor® 700 (clone 2H7) | BD Biosciences | Cat# 560631 |
| Mouse anti-human IgD APC-H7 (clone IA6-2) | BD Biosciences | Cat# 561305 |
| Mouse anti-human IgM BUV395 (clone G20-127) | BD Biosciences | Cat# 563903 |
| Mouse anti-human CD4 BV510 (clone SK3) | BD Biosciences | Cat# 562970 |
| Mouse anti-human CD16 BV510 (clone 3G8) | BD Biosciences | Cat# 563830 |
| Mouse anti-human CD10 BV650 (clone HI10a) | BD Biosciences | Cat# 563734 |
| Mouse anti-human IgG BUV496 (clone G18-145) | BD Biosciences | Cat# 741172 |
| Mouse anti-human CD14 BV510 (clone M5E2) | BioLegend | Cat# 301842 |
| Mouse anti-human CD3 BV510 (clone UCHT1) | BioLegend | Cat# 300448 |
| Mouse anti-human CD56 BV510 (clone 5.1H11) | BioLegend | Cat# 362534 |
| Mouse anti-human CD8a BV510 (clone RPA-T8) | BioLegend | Cat# 301048 |
| Mouse anti-human CD27 BV605 (clone O323) | BioLegend | Cat# 302830 |
| Mouse anti-human CD21 FITC (clone Bu32) | BioLegend | Cat# 354910 |
| Mouse anti-human CD38 PE/Cy7 (clone HIT2) | BioLegend | Cat# 303516 |
| Mouse anti-human CD19 PE/Dazzle 594 (clone HIB19) | BioLegend | Cat# 302252 |
| Mouse anti-human CD4 APC-H7 (clone SK3) | BD Biosciences | Cat# 641398 |
| Mouse anti-human CD8 PerCP (clone SK1) | BD Biosciences | Cat# 347314 |
| Mouse anti-human CD134 PE/Cy7 (clone ACT35) | BioLegend | Cat# 350012 |
| Mouse anti-human CD137 APC (clone 4B4-1) | BD Biosciences | Cat# 550890 |
| Mouse anti-human CD69 BV421 (clone FN50) | BD Biosciences | Cat# 562884 |
| Mouse anti-human CD25 PE (clone M-A251) | BD Biosciences | Cat# 555432 |
| Rat anti-human CXCR5 Alexa Fluor® 488 (clone RF8B2) | BD Biosciences | Cat# 558112 |
