## Supplementary material for "Informing the need for a SARS-CoV-2 booster based upon the immune response among young, healthy adults to variants circulating during late 2023": References

LINKS COVID REFERENCES

14. Pooley N, Abdool Karim SS, Combadière B, Ooi EE, Harris RC, El Guerche Seblain C, Kisomi M, Shaikh N. Durability of Vaccine-Induced and Natural Immunity Against COVID-19: A Narrative Review. Infect Dis Ther. 2023 Feb;12(2):367-387. doi: 10.1007/s40121-022-00753-2. Epub 2023 Jan 9. PMID: 36622633; PMCID: PMC9828372.
15. Crawford KHD, Dingens AS, Eguia R, Wolf CR, Wilcox N, Logue JK, et al. Dynamics of neutralizing antibody titers in the months after severe acute respiratory syndrome coronavirus 2 infection. J Infect Dis [Internet]. 2021 [cited 2022 Apr 6];223:197–205.
16. Centers for Disease Control and Prevention. CDC recommends updated COVID-19 vaccine for Fall/Winter virus season. CDC, September 12, 2023 (https://www.cdc.gov/media/releases/2023/p0912-COVID-19-Vaccine.html).
17. Rohde, R. https://asm.org/Articles/2023/October/Updated-COVID-19-Booster-XBB15-What-to-Know. American Society for Microbiology, October 26, 2023 (https://asm.org/Articles/2023/October/Updated-COVID-19-Booster-XBB15-What-to-Know).
18. El-Sadr WM, Vasan A, El-Mohandes A. Facing the new COVID-19 reality. N Engl J Med 2023; 388:385-387.
19. Lizewski RA, Sealfon RSG, Park SW et al. SARS-CoV-2 Outbreak Dynamics in an Isolated US Military Recruit Training Center With Rigorous Prevention Measures. Epidemiology. 2022 Nov 1;33(6):797-807. doi: 10.1097/EDE.0000000000001523. Epub 2022 Aug 5. PMID: 35944149; PMCID: PMC9531985.
20. US Department of Defense. DOD Rescinds COVID-19 Vaccination Mandate. January 10, 2023 (https://www.defense.gov/News/Releases/Release/Article/3264323/dod-rescinds-covid-19-vaccination-mandate/).
21. Wiysonge CS, Ndwandwe D, Ryan J et al. Vaccine hesitancy in the era of COVID-19: could lessons from the past help in divining the future? Hum Vaccin Immunother. 2022 Dec 31;18(1):1-3. doi: 10.1080/21645515.2021.1893062. Epub 2021 Mar 8. PMID: 33684019; PMCID: PMC8920215.
22. Beusekom, MV. 'A deadly societal force': A Q&A with author Dr. Peter Hotez on the anti-science movement. CIDRAP, October 5, 2023 (https://www.cidrap.umn.edu/anti-science/deadly-societal-force-qa-author-dr-peter-hotez-anti-science-movement).
23. Howard J. Despite rocky rollout, more than 7 million Americans have received updated Covid-19 vaccines, HHS says. CNN, October 7, 2023 (https://www.cnn.com/2023/10/12/health/hhs-covid-vaccine/index.html).
24. Wiemken, T.L., Khan, F., Puzniak, L. *et al.* Seasonal trends in COVID-19 cases, hospitalizations, and mortality in the United States and Europe. *Sci Rep* **13**, 3886 (2023). https://doi.org/10.1038/s41598-023-31057-1
25. National Institute of Allergy and Infectious Diseases. Standard Operating Procedure: Peripheral Blood Mononuclear Cells(PBMC) and Associated Plasma Collection. Division of Microbiology and Infectious Diseases, Version 6.0, September 30, 2021. https://www.niaid.nih.gov/sites/default/files/PBMC_SOP.pdf.
26. World Health Organization. COVID-19 Epidemiological Update - 24 November 2023. Emergency Situational Updates, Edition 161, November 24, 2023 (file:///C:/Users/Huy.Nguyen/Downloads/20231124_covid-19_epi_update_161.pdf).
